# Benchmarking and quality control for nanopore sequencing and feasibility of rapid genomics in New Zealand: validation phase at a single quaternary hospital

**DOI:** 10.1101/2024.06.13.24307636

**Authors:** Denis M. Nyaga, Peter Tsai, Clare Gebbie, Hui Hui Phua, Patrick Yap, Polona Le Quesne Stabej, Sophie Farrow, Jing Rong, Gergely Toldi, Eric Thorstensen, Zornitza Stark, Sebastian Lunke, Kimberley Gamet, Jodi Van Dyk, Mark Greenslade, Justin M. O’Sullivan

**Affiliations:** Liggins Institute, The University of Auckland, New Zealand; Molecular Medicine and Pathology, The University of Auckland, New Zealand; Genetic Health Service New Zealand-Northern Hub, Te Toka Tumai, Auckland; Starship Child Health, Te Whatu Ora Te Toka Tumai, Auckland, New Zealand; Victorian Clinical Genetics Services, Murdoch Children’s Research Institute, Parkville, Melbourne, Australia; Faculty of Medicine, Dentistry and Health Sciences, The University of Melbourne, Melbourne, Australia; Diagnostic Genetics, Department of Pathology and Laboratory Medicine, Te Toka Tumai, Auckland

**Author notes:** These authors contributed equally to this work (Denis M. Nyaga, Peter Tsai, Clare Gebbie, Hui Hui Phua, Patrick Yap).

## Abstract

Approximately 200 critically ill infants and children in New Zealand are in high-dependency neonatal/paediatric acute care at any given time, many with suspected genetic conditions, necessitating a scalable distributed solution for rapid genomic testing. We adopt the existing acute care genomics protocol of an accredited laboratory and established an expandable acute care clinical pipeline based around the Oxford Nanopore Technologies PromethION 2 solo system connected to a Bayesian AI-based clinical decision support tool (Fabric GEM™ software). In the establishment phase, we performed benchmarking using Global Alliance for Genomics and Health (GA4GH) benchmarking tools and Genome in a Bottle samples HG002-HG007. We evaluated single nucleotide variants (SNVs) and small insertions-deletions (indels) calls and achieved SNV precision and recall of 0.997 ± 0.0006 and 0.992 ± 0.001, respectively. Small indel identification approached a precision of 0.922 ± 0.019 and recall of 0.838 ± 0.043. Rarefaction analyses demonstrated that SNV identification plateaus at ∼20X coverage, while small indels plateaus at ∼40X coverage. Large genomic variations from Coriell Copy Number Variation Reference Panel 1 (CNVPANEL01) were reliably detected with ∼2M long reads. Finally, we present results obtained from ten trio samples that were processed through the pipeline validation phase, averaging a 5-day turnaround time, conducted in parallel with a clinically accredited short-read rapid genomic testing pipeline.

## Introduction

The rise of clinical genomic testing, utilizing exome and whole genome sequencing, has enabled the detection of genomic changes (*i.e.* single nucleotide variants [SNVs], small insertions and deletions [indels], copy number variants [CNVs], structural variants [SVs]), and elucidated the underlying genetic basis of rare Mendelian disorders and cancers.^1–7^ Over the past decade, the increasing adoption of genomic testing has generated substantial evidence supporting precision medicine.^2,4,6^ Such approaches have enabled molecular diagnoses for genetic disorders, guiding tailored medical interventions.^6–9^

Genomic testing has been established using cost-effective, short-read (150 bp in fragments) sequencing platforms from Illumina (*i.e.* NovaSeq, HiSeq, MiSeq, NextSeq), Thermo Fisher (Ion Torrent sequencer), and BGI (*i.e.* BGISEQ and MGISEQ). However, short-read sequencing has well recognised limitations.^10,11^ Firstly, it is difficult to uniquely align short reads to complex repetitive genomic regions involved in short tandem repeat (STR) expansion disorders (*e.g.* Fragile X syndrome and Huntington’s disease).^10,12^ Secondly, the requirement for PCR amplification in short-read sequencing may contribute artefacts and hinder the identification of native base modifications.^10^ Thirdly, short read lengths hinder the identification and precise phasing of alleles in large SVs.^10^

The recently developed long read sequencing (LRS) platforms (*i.e*. Oxford Nanopore Technologies [ONT] and Pacific Biosciences [PacBio]) employ direct inspection of single molecules during DNA synthesis, yielding long phaseable reads (>10 kb) in real time.^12–15^ Consequently, long reads generate highly reliable complete genome assemblies^15^, which can serve as benchmarks for short-read data. The utilization of ONT long reads as a standalone sequencing platform in clinical diagnosis has been demonstrated.^16–18^ In the research setting, LRS has been used to: a) identify and fine-map structural variations at single-nucleotide resolution; and b) resolve the haplotypes of heterozygous SVs.^13–14,19^ Novel pathogenic variants have been uncovered by LRS technology in human diseases with a previously unknown underlying genetic cause.^20^ Additionally, long reads facilitate the characterization of pathogenic repeat expansions in genomic regions that are challenging to sequence using short-read sequencing technology.^21–22^

The clinical application of LRS^13–22^ requires confidence in the accuracy of variant calling for SVs, CNVs, STRs, and SNVs/indels. However, high per-base error rates in low-complexity and homopolymer sequences^12,23^, and other issues have led to concerns about the application of ONT in clinical settings. Thus, there is a need for comprehensive benchmarking to: 1) confirm precision relative to the routinely used short-read technologies; and 2) illustrate the benefits and limitations of LRS technology for application to CGS.

The Global Alliance for Genomics and Health (GA4GH) developed benchmarking protocols to evaluate the performance of sequencing platforms and variant-calling methods before their integration into clinical practice.^24–26^ Benchmarking is essential to ensure adherence to standards and relies upon datasets where the relationship between input and output is known. This facilitates testing of consistency between the expected and observed outcomes (true positives).^24^ The Genome in a Bottle (GIAB) consortium offers reference samples (*e.g.* HG001 from the HapMap project and trios of Ashkenazi Jewish and Han Chinese ancestry from the Personal Genome Project) with ground-truth calls for SNVs, small indels, and SVs.^27,28^ Notably, GIAB recently provided a curated benchmark of challenging medically relevant genes through haplotype-resolved whole-genome assembly.^29^ The GA4GH resources enable performance assessment, optimization, and analytical validation of CGS assays and workflows for detecting genomic variations.^24,25^ Indeed, GIAB datasets and benchmarks are considered the gold standard for evaluating sequencing technologies and variant calling pipelines.^27^

The ONT platform generates sequencing data in real-time, allowing samples to be distributed across flow cells to reduce the sequencing time, where each additional flow cell reduces the sequencing time needed on a sample by 1/n (where n is the number of flow cells). Notably, this has been demonstrated in a recent study that sequenced a single human genome across 48 flow cells, generating high-depth genome-wide data (200 Gigabases) and candidate variant identification in less than eight hours.^30^ The ONT platform is also capable of targeted sequencing through adaptive sampling, which removes the need to design custom probes to capture genes or regions of interest through a dynamic and modifiable process during the sequencing run.^31^ DNA and RNA base modifications, including 5-methylcytosine (5mC), 5-hydroxymethylcytosine (5hmC), and N^6^-methyladenine (6mA), can also be detected computationally on raw ONT data without the need to perform special library preparations such as bisulphite conversion,^32^ which is known to cause DNA damage and can lead to overestimation of the 5mC level.^33^

We have established an expandable rapid genomic testing pipeline based around the ONT PromethION2 (P2) solo system connected to AI-driven genomics analysis and interpretation software (*i.e.* Fabric GEM™ software) for tertiary analysis. In the establishment phase, we benchmarked our pipeline using GAG4H tools and GIAB reference cell lines HG002 - HG007 for SNVs and small indels analysis. In addition, we used CNVPANEL01 (Coriell Institute) to measure our ability to detect large-scale chromosomal abnormalities. Finally, we present the results of the pipeline validation phase, performed in parallel with a clinically accredited short-read rapid genomic testing service.

## 3. Methods

### 3.1. Benchmarking of the sequencing platform and variant-calling methods

#### 3.1.1. Samples and truth sets

We acquired CNVPANEL01 as 3 µg genomic DNA (at 100 µg/ml) per sample and GIAB reference samples (*i.e.* HG002 - HG007) with available truth sets, from the Coriell repository (Coriell Institute for Medical Research, 403 Haddon Avenue Camden, NJ 08103, USA).

#### 3.1.2. Library preparation and sequencing

DNA samples (1500 ng) were sheared to 10-15kb using Covaris g-TUBES (Covaris) in a bench-top centrifuge for 1 minute at 2000 RCF (room temperature). Nanopore sequencing libraries were prepared according to the genomic DNA Ligation Sequencing Kit V14 (SQK-LSK114) protocol (ONT, Oxford Science Park, OX4 4DQ, UK). Prepared libraries were loaded on PromethION flow cells (R10.4) and sequenced (*i.e.* depth of between 24-42X) with the PromethION 2 (P2) solo device using Kit 14 chemistry and MinKNOW v23.07.8 (Oxford Nanopore Technologies [ONT], Oxford Science Park, OX4 4DQ, UK).

#### 3.1.3. Read base calling and variant calling

Base calling of raw ONT signal data was completed using Dorado v0.3.3 (https://github.com/nanoporetech/dorado) with the high accuracy (hac) model (dna_r10.4.1_e8.2_400bps_hac@v4.2.0). In addition, base calling of the HG002 sample was also completed with the super accuracy (sup) model (dna_r10.4.1_e8.2_400bps_sup@v4.2.0). The resulting FASTQ files, with a Phred quality score (Q score) >9, in the fastq_pass folder, were processed with EPI2ME Labs’ wf-alignment pipeline (https://github.com/epi2me-labs/wf-alignment; v0.5.2). Briefly, FASTQ files were aligned to the GRCh38 reference genome using minimap2 (v2.26).^34^ EPI2ME Labs’ wf-human-variation pipeline (https://github.com/epi2me-labs/wf-human-variation; v1.7.0) was subsequently employed for genomic variant processing, including SNV and small indel calling with Clair3 (v1.0.4)^35^, SV calling with Sniffles2 (v2.2)^36^, and CNV calling with QDNAseq (v1.38)^37^ using default parameters, with a VNTR annotation file provided for accurate SV identification. Repeat expansions were genotyped using Straglr (https://github.com/philres/straglr)^38^ as implemented in EPI2ME Labs’ wf-human-variation pipeline v1.7.0.

#### 3.1.4. Benchmarking of variant calling

Variant comparison tools (https://github.com/ga4gh/benchmarking-tools)^24^ are integral to genomic benchmarking as they identify shared variations between ground-truth calls and comparison results (*i.e*. true positives [TP]), along with variants unique to each set (*i.e*. false negatives [FN]), and additional variants (*i.e*. false positives [FP]). We compared called SNVs and small indels with GIAB ground-truth variants (benchmark version v4.2.1)^24^ using hap.py v0.3.15 (https://github.com/Illumina/hap.py), and each variant was labelled as TP, FP, or FN. Hap.py also provides precision (positive predictive value [PPV]), recall (sensitivity) and F1 scores (harmonic mean of precision and recall) calculated as follows:

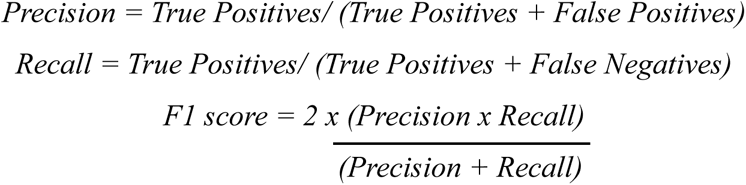

For SVs, we employed Truvari v4.1.0 (https://github.com/ACEnglish/truvari)^39^ to benchmark variants with GIAB ground-truth SVs. Each variant was categorized as TP, FP, or FN based on this comparison.

#### 3.1.5. Rarefaction analysis

Rarefaction was performed to evaluate the sensitivity and reliability of long read variant calling across different sequencing depths. Subsampling of the Binary Alignment Map (BAM) files was performed using Samtools^40^, by randomly selecting subsets of reads from the original alignment files. The subsampled BAMs were then subjected to variant calling analysis as described (Section 3.3). Benchmarking for SNVs and small indels was conducted as detailed (Section 3.4). Rarefaction curves were generated using python v3.10.8 and the seaborn v0.12.2 library to illustrate the relationship between sequencing depth and the called variants, enabling the evaluation of variant calling performance and reliability across varied sequencing depths.

#### 3.1.6. Benchmarking analysis for challenging clinically relevant genes

We called SNVs and small indels across genomic regions overlapping challenging clinically relevant genes^29^ using the original BAM files and pipeline outlined in Section 3.3. Benchmarking for SNVs and small indels was conducted as detailed (Section 3.4).

#### 3.1.7. Methylation analysis

Raw ONT signal data in POD5 files (https://github.com/nanoporetech/pod5-file-format) was base called (Dorado v0.5.0) using the high accuracy (hac) DNA base modification model (dna_r10.4.1_e8.2_400bps_hac@v4.2.0_5mCG_5hmCG@v2) to detect modified bases (*i.e.* 5- methylcytosine [5mC], 5-hydroxymethylcytosine [5hmC]). The modified BAM files (modBAMs) were aligned to the GRCh38 reference genome and modkit v0.2.3 (https://github.com/nanoporetech/modkit) employed to generate genome-wide summary counts of modified and unmodified bases into bedMethyl files.

#### 3.1.8. Visualization

Plots were generated using the seaborn v0.12.2 and matplotlib v3.7.1, and python v3.10.8.

### 3.2. Newborn Genomics Programme (NBG) protocol for patient recruitment, sample collection, DNA extraction, sequencing and variant analysis

#### 3.2.1. Design

This is a research study to determine the medical and economic impacts of rapid whole genome sequencing (rWGS) within the New Zealand health care landscape. Ethics approval was obtained from the Northern B Health and Disability Ethics Committee for the study entitled: *Newborn Genomics – Te Ira oo Te Arai* (Ethics reference: 2023 FULL 15542). Locality approval was obtained from the Research Review Committee Te Toka Tumai Auckland for the project entitled: *Newborn Genomics – Te Ira oo Te Arai* (Reference A+9855 [FULL 15542]). This study is registered in ClinicalTrials.gov (NCT06081075). The clinical protocol was adopted and modified as per Lunke *et al*., 2023.^41^

#### 3.2.2. Participants

Children with suspected genetic conditions and their families were recruited into the study from the neonatal and paediatric intensive care units (*i.e*. NICU and PICU, respectively) and the National Metabolic Service at Te Toka Tumai | Auckland City Hospital. Within NICU, participation was limited to proband-parent trios of critically sick neonates with evidence of a suspected genetic condition, without a clear non-genetic aetiology, or who developed an abnormal response to standard therapy for an underlying condition within the preceding seven days. For infants within PICU or under the care of Metabolic Services, participation was limited to proband/parent trios of children with an acute or chronic illness with evidence of a suspected genetic condition without a clear non-genetic aetiology.

All participants continued to receive the standard of care, irrespective of whether they were included in the study.

#### 3.2.3. Referral of participants to the study

Potential participants were referred to the geneticist on-call (by telephone) for a formal genetic review, mainly by a neonatologist or a paediatric intensivist or the lead paediatric subspecialist for the patient when a genetic condition was suspected, or when the aetiology of a condition was unclear and a genetic cause needed to be ruled out to guide further clinical management.

#### 3.2.4. Inclusion and exclusion criteria

Inclusion and exclusion criteria were modified from Dimmock *et al*.^42^ and McKeown *et al.*^43^

The inclusion criteria was:

- acutely ill inpatient
- admitted to NICU or PICU between April 2023 – March 2026
- under the care of the National Metabolic Service between April 2023 – March 2026
- within 1 week of hospitalization or within 1 week of developing abnormal response to standard therapy for an underlying condition
- suspected genetic condition, without a clear non-genetic aetiology

The exclusion criteria was:

- patients whose clinical course is entirely explained by

- isolated prematurity
- isolated unconjugated hyperbilirubinemia
- infection or sepsis with expected response to therapy
- a previously confirmed genetic diagnosis that explains the clinical condition
- isolated transient neonatal tachypnoea
- meconium aspiration syndrome
- trauma
- inability to source blood or buccal samples for DNA extraction from at least the mother and child

Of note, participants were only considered for the study if they were referred to clinical genetics as a part of their standard of care workup.

#### 3.2.5. Recruitment multidisciplinary meeting

Following the referral of potential participants to the study, a multidisciplinary meeting (MDM) was convened via video conference to evaluate the eligibility of the referral based on the study’s inclusion and exclusion criteria. At a minimum, the MDM was comprised of a clinical geneticist, genetic counsellor, principal investigator, project manager, representative from the genomic analytical team (bioinformatician, variant curator) and the referring clinician. After agreeing to participate, the patients were registered on RedCap, and a study reference was generated. Subsequently, the clinical geneticists and genetic counsellors completed the clinical information, including phenotypic characterization using HPO terms, and facilitated informed consent.

#### 3.2.6. Pretest counselling and informed consent

Parents or guardians of the proposed probands were informed of the details of the study using the HDEC-approved Newborn Genomics Programme Participant Information Sheet (**Supplementary File 1**), and had the opportunity to ask questions to an on-call geneticist and genetic counsellor from the Genetic Health Service New Zealand. Written informed consent was obtained from parents or guardians before any study specific processes were undertaken (Newborn Genomics Programme consent form, **Supplementary File 2**).

#### 3.2.7. Phenotyping

Clinical geneticists and subspecialists performed clinical phenotyping, which was recorded on RedCap using the Human Phenotype Ontology (HPO) terms (https://hpo.jax.org/app/) to optimize phenotypic data exchange during the curation stages of the analysis. At the same time, a phenotype-focus gene list was generated using PanelApp (Australia [https://panelapp.agha.umccr.org/] and the UK [https://panelapp.genomicsengland.co.uk/]) and shared with the genomic analytical team for inclusion in the Bayesian AI-based clinical decision support tool (Fabric GEM™ software).

#### 3.2.8. Sample collection

After obtaining consent, duplicate blood samples were collected: 4 mL EDTA blood samples from the mother and father, and 500 µL EDTA blood samples from the child. One set of samples was sent to the Liggins Institute newborn genomics laboratory for sequencing and variant analysis, while the second set was sent to the clinical laboratory, Victorian Clinical Genetics Services (VCGS) in Melbourne, Australia, for a concurrent, independent, short-read-based analysis as described in Lunke *et al*., 2023.^43^

#### 3.2.9. DNA extraction, library preparation, sequencing, and variant calling

High molecular weight DNA was extracted from 300µl of the whole blood using the Puregene DNA extraction Kit (Qiagen) following the manufacturer’s protocol, and the extracted DNA eluted in nuclease-free water (Thermo Fisher Scientific). The quantification and purity assessment of the DNA samples were performed using the Qubit system (Thermo Fisher Scientific) and a spectrophotometer (Implen NanoPhotometer). The library preparation and sequencing procedures were carried out as detailed in section 3.1.2. Finally, the base calling of sequenced reads and variant calling analysis was conducted following the methods described in section 3.1.3.

#### 3.2.10. Variant review multi-disciplinary meeting

Genomic variants were prioritized using Fabric GEM™ software after the data was generated and annotated following standard protocols. These prioritized variants were manually curated by variant curators. A multidisciplinary review meeting (MDM) was then held to evaluate the results. The review MDM comprised the same clinicians and study representatives who attended the recruitment MDM. During the meeting, the genomic data analysts presented the quality control report and discussed the prioritized variants, and the evidence for pathogenic/likely pathogenic variants, for genotype-phenotype correlation. The VCGS results were not shared with the NBG team, ensuring they remained blinded to the clinically validated results until the variant review MDM. Finally, the clinical geneticist and genetic counsellor disclosed and discussed the molecular diagnosis based on the accredited acute care genomics service (VCGS) results.

#### 3.2.11. Genomic reports

Upon completing the variant review meeting, the NBG study team generated a research report. Simultaneously, the clinical laboratory (VCGS) produced its validated clinical report, which was directly returned to the clinical team for disclosure to the families. Finally, the genetic counsellor communicated the study report findings to the study participants participant and addressed any discrepancies identified in the reports.

## 4. Results

### 4.1. Overview of genomic benchmarking workflow for acute care clinical pipeline

We have performed genomic variant benchmarking of an expandable acute care clinical pipeline, using the set standards and guidelines provided by GAG4H.^24^ DNA samples from the Coriell repository, including the characterized GIAB reference samples (HG002 - HG007) with available truth sets, were used for benchmarking variant calling of SNVs (*i.e.* single base substitutions) and small indels (*i.e.* insertions and deletions <50 bps) (**Figure 1**). Sample HG002 was used to benchmark SVs (*i.e.* genomic alteration >50 bps encompassing insertions, deletions, duplications, inversions, and translocations). Coriell samples carrying pathogenic variants (*i.e.* GM06936, GM06870, GM01416, GM20556, GM09367, GM05966, GM05067, GM09216) were used to evaluate the performance of long reads in the identification of large-scale chromosomal abnormalities (**Figure 1**).

**Figure 1.**
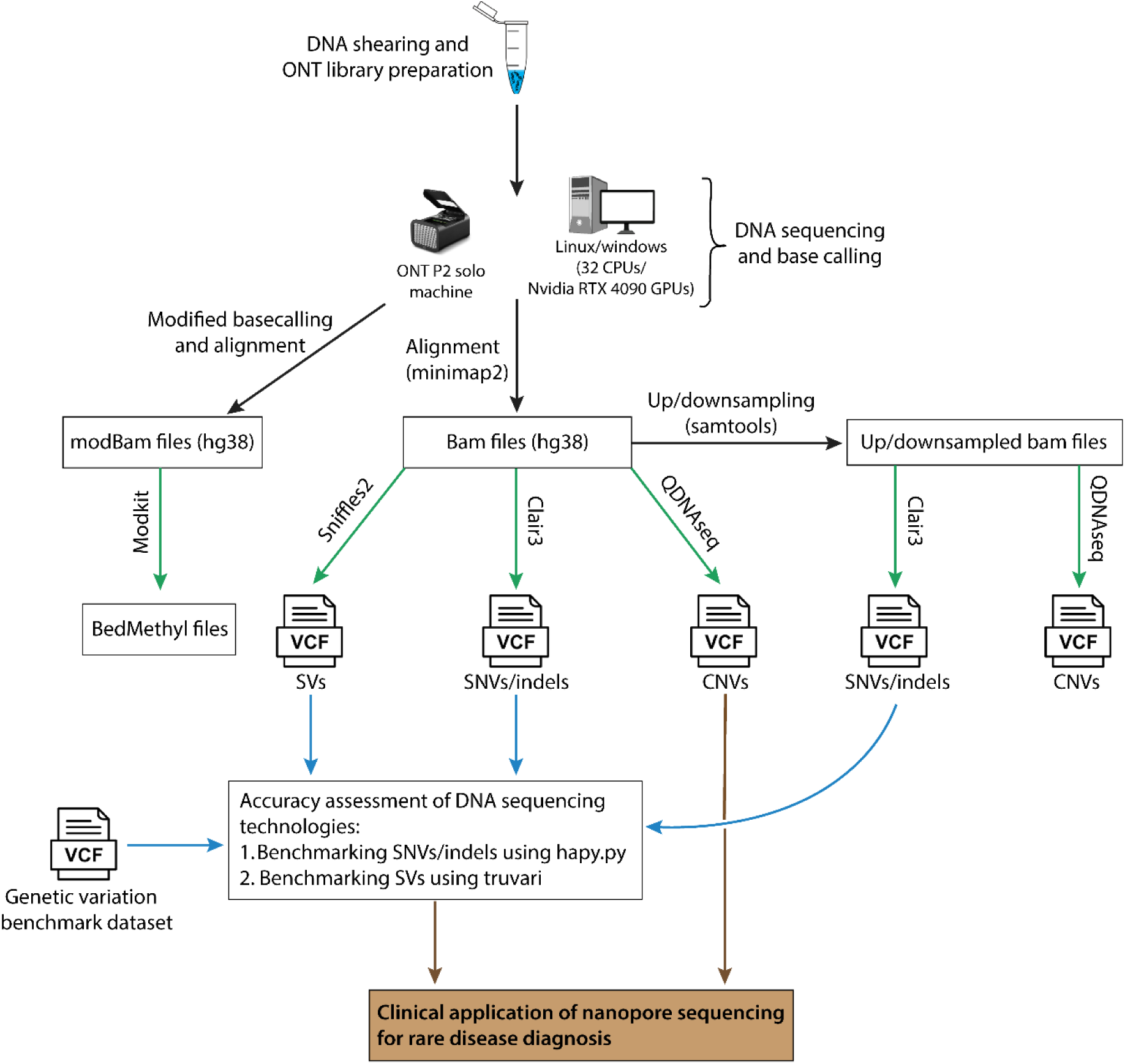
An overview of the genomic benchmarking for the acute care clinical pipeline. Genomic DNA sequencing and variant calling was performed using GIAB cell lines (HG002 - HG007) and Coriell Institute for Medical Research CNVPANEL01 cell lines. ONT sequencing libraries were prepared and sequenced using PromethION flow cells (R10.4). Variant calling was performed using EPI2ME pipeline that includes: a) clair3 for SNVs and small indels analysis, b) sniffles2 for SVs calling; and c) QDNAseq for CNVs analysis. SNVs and small indels were benchmarked against GIAB ground-truth variants using hap.py v0.3.15 (NIST v4.2.1), and Truvari v4.1.0 was employed to benchmark SVs (NIST v0.6). Modified base calling and alignment to GRCh38 reference genome was performed using dorado v.5.0 and genome-wide summary counts of 5-methylcytosine (5mC) and 5-hydroxymethylcytosine (5hmC) were generated using modkit v0.2.3.

### 4.2. ONT long reads provide high precision for small variant calls

Following sequencing using the R10.4 pore, base calling, and variant calling, we obtained mean read lengths of 4.2 kb, mean alignment accuracy of 97.2%, read N50 of 7.6 kb, and average depth of coverage of 37.9X for six samples (**Figure 2A**). Benchmarking was performed to evaluate the general performance of ONT reads on SNVs and small indels (up to 50 bps) calling from GIAB samples HG002 – HG007 (see Methods). Precision, recall, and F1 scores were computed against truth sets (National Institute of Standards and Technology [NIST)] benchmark v4.2.1; https://github.com/ga4gh/benchmarking-tools/blob/master/resources/high-confidence-sets/giab.md), for: 1) high-confidence regions excluding homopolymers, defined as four or more consecutive identical nucleotides ±1 base pair on each side; 2) genome-wide coding regions, including the Mendeliome; and 3) 273 challenging medically relevant genes for the HG002 genome (CMRG v1.0; https://data.nist.gov/od/id/mds2-2475).

**Figure 2.**
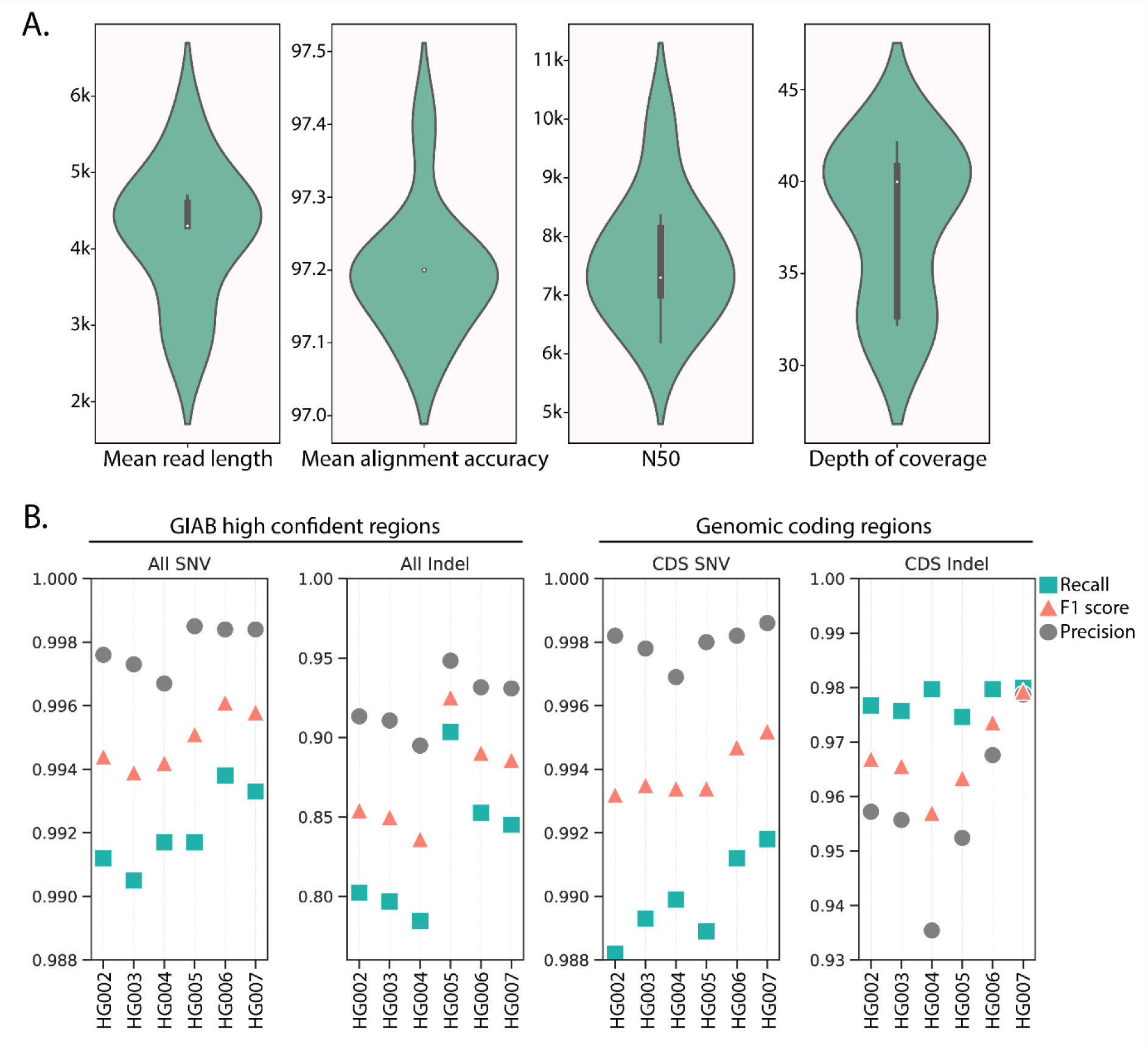
Sequencing metrics and variant calling accuracy ONT summary statistics. A) Violin plots showing mean read length, mean alignment accuracy (%), N50, and mean depth of coverage for GIAB samples. **B)** Comparisons of precision (positive predictive value [PPV]), recall (sensitivity) and F1 scores (harmonic mean of precision and recall) for SNVs and small indels called from ONT LRS data compared to GIAB high-confidence regions (left) and coding regions excluding homopolymers and difficult-to-map genomic regions (right) for GIAB samples HG002-HG007. A summary of benchmarking metrics across two separate sequencing runs for HG002-HG007 samples is available as **Supplementary Table 1**. CDS: coding sequence; N50: the length of the shortest read among the longest sequences, encompassing ∼50% of the total nucleotides in a set of sequences.

Across two separate sequencing runs, the average SNV precision and recall were 0.998 and 0.992), respectively, while small indel precision and recall were 0.922 and 0.831, respectively, within GIAB high-confidence regions (**Figure 2B; Supplementary Table 1**). When assessing variants exclusively within coding regions and regions excluding homopolymers and difficult-to-map genomic regions, small indels achieved precision >0.935 and recall >0.974 (**Figure 2B; Supplementary Table 1**). Furthermore, we assessed the performance of long reads on identifying variants in CMRG. ONT LRS demonstrated precision and recall scores >0.967 and >0.978, respectively, for SNVs and >0.836 and >0.701, respectively, for small indels within the 273 genes in the CMRG set (**Table 1**). We observed slightly improved precision and recall metrics for small indels called from the super accuracy (sup) base called HG002 genome (*i.e.* precision = +0.042, recall = +0.011 for high-confidence regions; and precision = +0.03, recall = +0.001 for the CMRG set; **Supplementary Table 2**). Overall, these results are consistent with previous benchmarking reports^44^ and validate the efficacy of the EPI2ME Labs’ implementation of Clair3^35^ in generating high-quality small variant calls comparable to gold-standard results.

**Table 1.** Benchmarking metrics for SNVs and small indels within CMRG.

| GIAB Sample | Run | SNP |  |  |  |  |  |  | INDEL |  |  |  |  |  |  |
| --- | --- | --- | --- | --- | --- | --- | --- | --- | --- | --- | --- | --- | --- | --- | --- |
|  |  | Truth Total | TP | FN | FP | Recall | Precision | F1 | Truth Total | TP | FN | FP | Recall | Precision | F1 |
| HG002 | 1 | 17,559 | 17,192 | 367 | 579 | 0.979 | 0.967 | 0.973 | 3,605 | 2,538 | 1,067 | 507 | 0.704 | 0.840 | 0.766 |
|  | 2 | 17,559 | 17,165 | 394 | 540 | 0.978 | 0.969 | 0.974 | 3,605 | 2,528 | 1,077 | 520 | 0.701 | 0.836 | 0.763 |

TP, true positive; FP, false positive; FN, false negative. Precision (positive predictive value [PPV]), recall (sensitivity) and F1 scores (harmonic mean of precision and recall).

### 4.3. Increasing sequencing depth beyond 40X does not improve small variant detection

Sequencing depth has been identified as being critical for the accurate identification of variants for the diagnosis of genetic diseases.^45^ We determined the optimal genomic depth for precise small variant identification from ONT reads. We downsampled the HG005 alignment file (∼40X) by randomly extracting sets of reads (*i.e.* at proportions of 0.12, 0.25, 0.3, 0.4, 0.5, 0.6, and 0.75 of the total set) to simulate sequence data of the same sample at sequencing depths of 4.8X, 10X, 12X, 16X, 20X, 24X, and 30X. Upsampling was performed at proportions of 1.25, 1.5, 1.75, and 2 to mimic depths of 50X, 60X, 70X, and 80X. Our analysis revealed that beyond a sequencing depth of 40X, there were no significant improvements in the detection of SNVs and small indels (**Figure 3**). These findings indicate that ∼40X is the optimal depth for accurate small variant discovery from ONT reads, and additional depth beyond this threshold does not enhance the accuracy or sensitivity of small variant detection.

**Figure 3.**
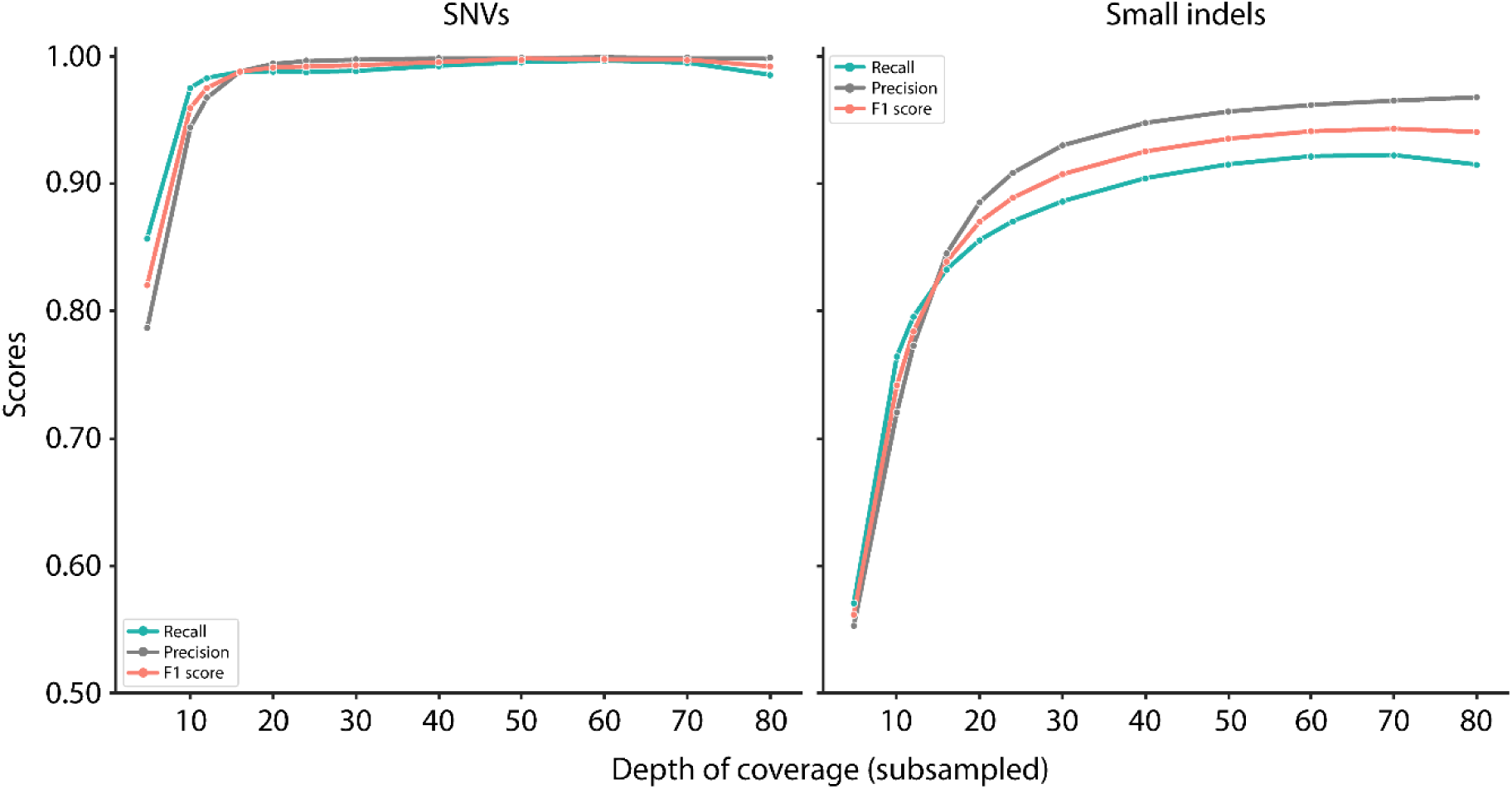
Sequencing depth exceeding 40X does not improve SNV or small indels detection. Rarefaction analysis of subsampled HG005 BAM files reveal consistent precision and recall scores for SNVs at 40X and 80X, with scores consistently at 0.996 and 0.998, respectively (left panel). Similarly, precision and recall scores for small indels (<50 bp) vary by <4% (*i.e*. 0.921 and 0.961, respectively) at depths of 40X and 80X (right panel).

### 4.4. ONT reads accurately identify structural variations and haplotype-specific tandem repeats

SVs were identified from the HG002 genome using Sniffles2 with a tandem repeat bed file provided to improve SV calling in repetitive regions (Methods).^36^ SV calling performance from two separate sequencing runs, together with publicly available ONT SVs (https://labs.epi2me.io/giab-2023.05/), were benchmarked against the publicly available high-confidence GIAB ground-truth SVs (from GRCh37 reference genome) and SVs called within the CMRG genes using Truvari v4.1.0. ONT reads demonstrated high precision (>0.951) and recall (>0.943) for high-confident regions (**Table 2**). Additionally, we observed precision and recall metrics of >0.889 and 0.946, respectively, for SVs within CMRG genes (**Table 3**), consistent with published benchmarks on ONT long reads.^44^

**Table 2.** Performance metrics for identification and genotyping of high-confident SVs.

| Sample | SV |  |  |  |  |  |
| --- | --- | --- | --- | --- | --- | --- |
|  | TP | FN | FP | Recall | Precision | F1 score |
| HG002 run 1 | 9,094 | 547 | 455 | 0.943 | 0.952 | 0.948 |
| HG002 run 2 | 9,234 | 407 | 474 | 0.958 | 0.951 | 0.954 |
| ONT public data | 9,396 | 245 | 486 | 0.975 | 0.951 | 0.963 |

**Table 3.** Performance metrics for identification and genotyping SVs within CMRG genes.

| Sample | SV |  |  |  |  |  |
| --- | --- | --- | --- | --- | --- | --- |
|  | TP | FN | FP | Recall | Precision | F1 score |
| HG002 run 1 | 196 | 10 | 11 | 0.907 | 0.947 | 0.927 |
| HG002 run 2 | 176 | 24 | 11 | 0.889 | 0.946 | 0.916 |
| ONT public data | 197 | 19 | 9 | 0.912 | 0.956 | 0.934 |

We profiled genomic regions associated with repeat expansions since these regions contribute to the development of numerous neurodevelopmental disorders (NDDs) (*e.g*. congenital and childhood-onset myotonic dystrophy type 1^46^). Using Straglr,^38^ we genotyped and quantified 37 clinically relevant tandem repeat regions (including *DMPK* and *NOTCH2NLC*) in the HG002 genome. Our findings indicate that long reads enable the genotyping of these regions (**Supplementary Figure 1**), while also providing haplotype-specific information for the repeat elements (**Supplementary** Figure 2).

### 4.5. Accurate identification of copy number variation at 2X sequencing depth

CNVs occur in neonatal disorders (*e.g*. 22q11.2 deletion syndrome^47^). In clinical testing, whole genome sequencing is poised to replace chromosomal microarray for the detection of CNVs.^45^ As such, the clinical utility of long read sequencing in identifying CNVs has been demonstrated.^17^ We assessed whether our variant calling workflow accurately detects clinically relevant pathogenic large chromosomal aberrations from Coriell samples (**Table 4**). Benchmarking results confirmed the reliable and accurate detection of these pathogenic CNVs using long-read sequencing (**Table 4**). Notably, we successfully identified two CNVs at ∼2X coverage by downsampling the BAM files to ∼2.6M reads: 1) the isodicentric chromosome CNV (*i.e*. 47,XY,+idic(15)(q13).ish idic(15)(q13)(D15Z1++,D15S11++,GABRB3++).arr Yq11.223q11.23(23920264-27079691)x2,15q11.1q13.3(18276329-30557740)x4); and 2) the XXXX syndrome CNV (*i.e*. 48,XXXX). (**Supplementary Figure 3**). Collectively, these results underscore the potential for long-read genomic testing in neonatal intensive care for diagnosis of suspected genetic conditions resulting from large chromosomal events.

**Table 4.**
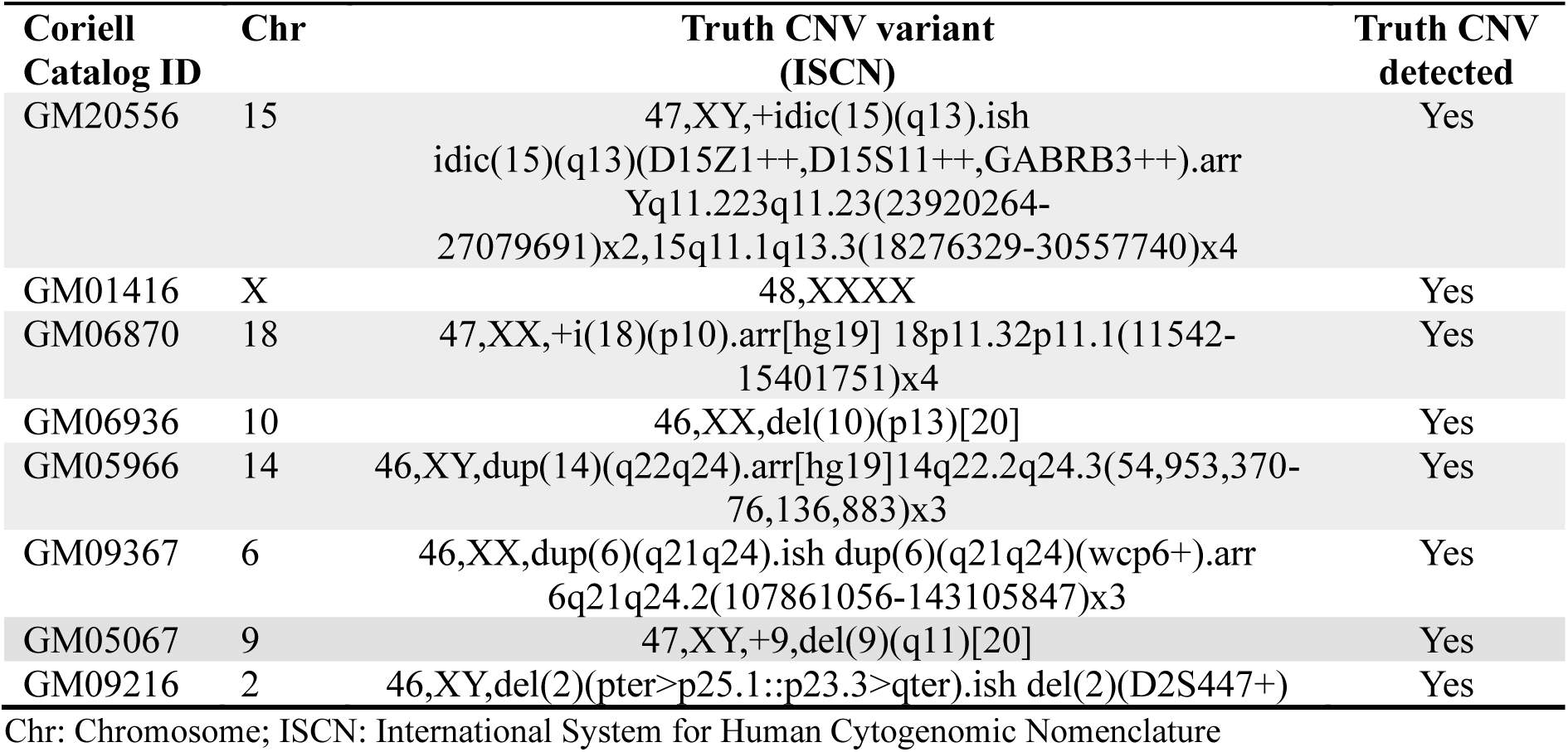
Long reads accurately detect pathogenic CNVs from CNVPANEL01 samples.

### 4.6. High concordance between ONT methylation calls and bisulphite sequencing

DNA methylation (5mC) is implicated in the pathogenic mechanism of *FMR1*-related disorders (*e.g*. fragile X syndrome), with expanded alleles typically exhibiting promoter hypermethylation and silencing of *FMR1*.^48^ As such, DNA methylation profiling is essential for complete genetic diagnosis of *FMR1*-related disorders.^48^ Notably, ONT sequencing facilitates concurrent profiling and quantification of DNA methylation (5-methylcytosine, 5mC). Genome-wide 5mC characterization of the HG002 genome using ONT sequencing identified 28.8 million CpG sites (98% of total GRCh38 CpG sites). Comparing ONT methylation calls to standard whole-genome bisulphite sequencing (WGBS) of the HG002 genome, acquired from the ONT open data repository (https://labs.epi2me.io/gm24385-5mc), identified a strong correlation (r=0.949; **Figure 5A**), consistent with highly accurate methylation calling. Notably, we identified haplotype-specific differentially methylated regions (DMRs) within gene promoters for imprinted genes (**Figure 5B**), as well as in novel DMRs (**Supplementary Figure 4**). This illustrates the potential utility of the haplotype-level resolution offered by ONT-based sequencing reads.

**Figure 5.**
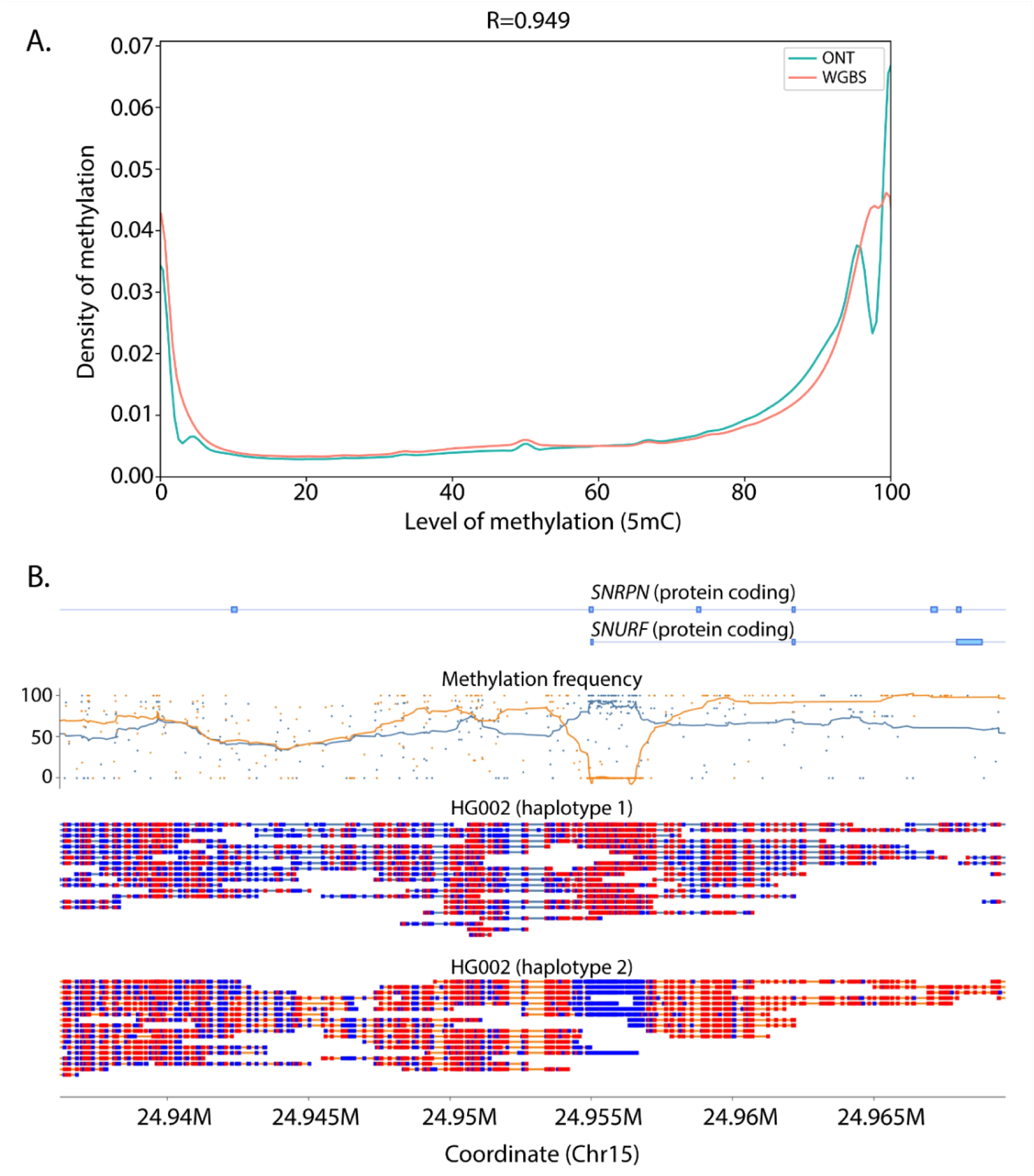
Strong correlation between 5mC methylation calls from ONT and whole-genome bisulphite sequencing. **A)** Density plots of the level of 5mC base modification across CpG sites detected through ONT (green) and bisulphite sequencing (orange) from the HG002 sample. Pearson correlation analysis identified a strong correlation for methylation levels across CpG sites between the ONT and whole genome bisulphite sequencing technologies. **B)** The *SNURF-SNRPN* locus exhibits haplotype-specific differential methylation, linked to maternal imprinting (paternal-expressed allele) as visualized using modbamtools (https://github.com/rrazaghi/modbamtools). Methylated CpG sites are denoted in red, while unmethylated sites are represented in blue. A methylation frequency plot and gene locus are visualized above the haplotype reads.

### 4.7. Application of the long-read pipeline in acute care genomic diagnosis

Our goal was to develop a scalable acute care genetic diagnostic pipeline by harnessing the capabilities of the ONT PromethION 2 solo system integrated with Fabric GEM™ (an AI-driven genomics analysis and interpretation software; https://fabricgenomics.com/). This integration was designed to facilitate precise genome annotation with rapid variant interpretation and prioritisation. The ultimate goal was to provide clinicians with access to actionable information pertaining to SNVs, small indels, and SVs **(Figure 6).**

**Figure 6.**
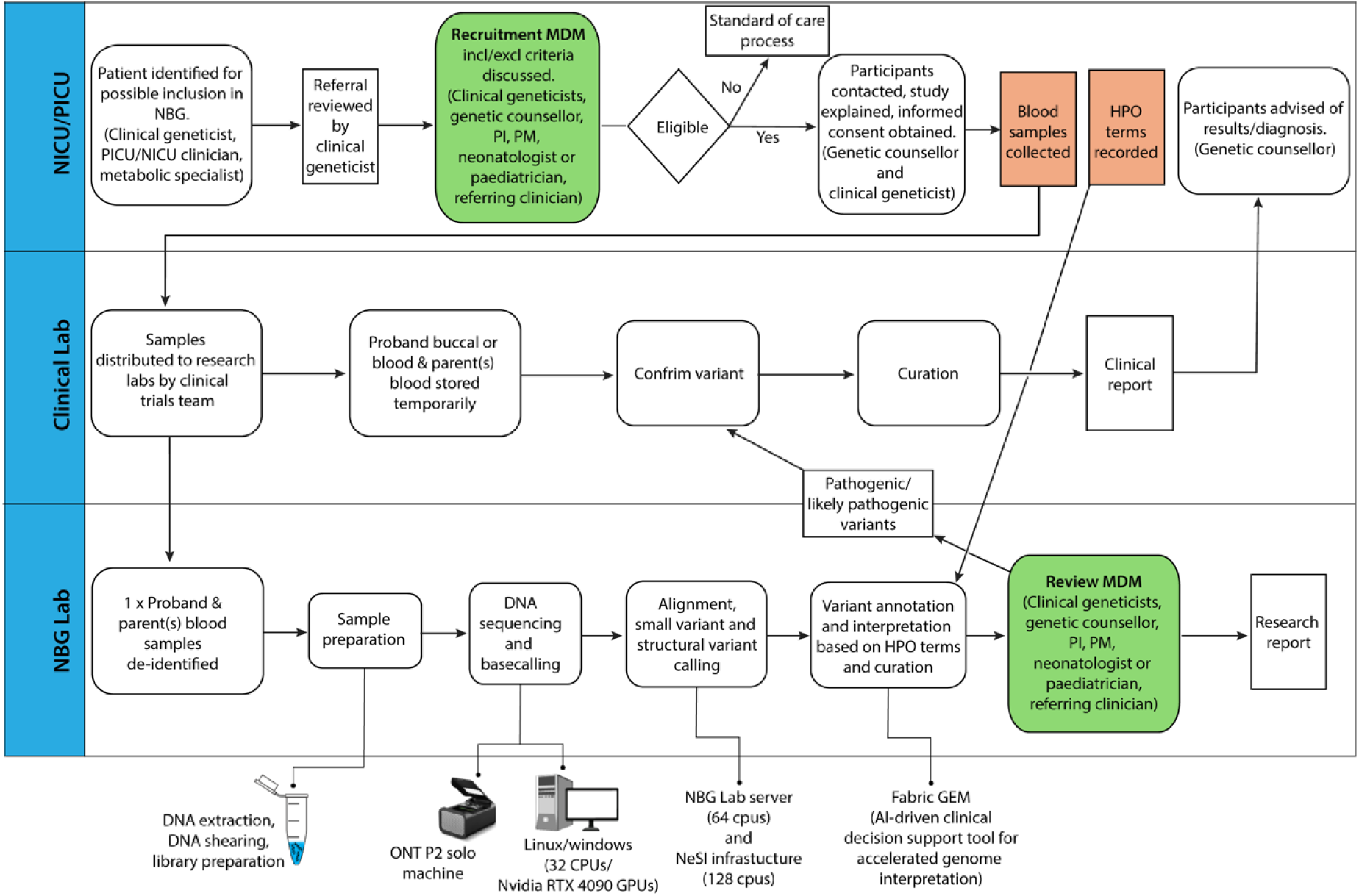
Flowchart for a scalable acute care clinical pipeline for rapid genome sequencing, precise genome annotation and accelerated variant interpretation using Human Phenotype Ontology (HPO) terms. This pipeline serves the purpose of informing acute management strategies for critically ill newborns, infants, and children suspected to have genetic disorders (see section 3.2). The process begins with the recruitment of patients admitted to the neonatal and paediatric intensive care (NICU and PICU) (in this instance at Starship Child Health) into the Newborn Genomics programme (NBG) following recommendations from a multidisciplinary team comprising clinical geneticists, genetic counsellors, neonatologists, paediatricians, and referring clinicians. Once consented, blood samples (from mother, father) and a buccal or blood sample (from the child) are collected in duplicate. One set of samples is sent to the NBG laboratory for sequencing, and the second to the clinical laboratory (in this instance at the Victorian Clinical Genetics Services [VCGS] in Melbourne, Australia) for variant confirmation. Samples sent to the NBG lab are sequenced using the ONT PromethION 2 solo system. Subsequently, sequence alignment and variant calling is performed using established infrastructure (*i.e.* EPI2ME Labs’ pipelines, including alignment of genomes to the GRCh38 reference genome with minimap2 [v2.26]^34^, SNV and small indel calling with Clair3 [v1.0.4]^35^, SV calling with Sniffles2 [v2.2]^36^, and CNV calling with QDNAseq [v1.38]^37^), with precise genome annotation and rapid variant interpretation conducted using Fabric GEM™. A further multidisciplinary meeting assesses the evidence for the identified variants with the patient’s phenotype. Orthogonal confirmation (*i.e.* Sanger Sequencing) of the candidate genetic variants is performed by accredited genetic testing facilities (in this instance at VCGS). Finally, clinical (from accredited genetic testing facilities) and research (from NBG) reports are generated, summarizing the evidence for the identified variants, with the clinical report forwarded to the genetic counsellor for communication of the genetic diagnosis to the participants.

During the establishment phase of this pipeline, ten critically sick children in the neonatal and paediatric intensive care (NICU and PICU) at Te Toka Tumai/Starship Child Health were referred for rapid long-read genomic sequencing (Ethics: Approval from Health and Disability Ethics Committee [Reference 2023 FULL 15542]; Locality approval: A+ 9855 [FULL15542]). In parallel, samples were provided to a clinically accredited genomics laboratory (VCGS; Melbourne, Australia) for rapid short-read Illumina genomic sequencing as described in Lunke *et al*., 2023.^43^ Sequencing, genome variant curation and analysis were independently undertaken at each site. Finally, genomic results from the accredited laboratory were independently provided to the clinician so that the long-read provider was unaware of the accredited results until after their results were presented to the MDM. Identical results were obtained across the ten proband-parents trios that were entered into the programme (**Table 5**). The identification of identical genomic findings demonstrates the applicability and reliability of our pipeline for acute-clinical care.

**Table 5.** Clinical characterisation and genomic results from ten proband-parent trio genome sequencing using short-read sequencing and long-read Nanopore sequencing.

| Family | Genomic results<br>(Liggins NBG - Acute care<br>genomic research) |  | Genomic results<br>(VCGS –Acute care<br>genomic service) |  | Variant<br>type/mode<br>of<br>inheritance | Clinical diagnosis<br>(phenotypic<br>characterisation in<br>proband(s)) | Clinical<br>indication<br>& Outcome | Turn-<br>around<br>time<br>(TAT;<br>days)* |
| --- | --- | --- | --- | --- | --- | --- | --- | --- |
|  | Gene | Variant<br>(classification) | Gene | Variant<br>(classification) |  |  |  |  |
| 1 | *KLHL40* | NM_152393.4:<br>c.1516A>C<br>p.(Thr506Pro)<br>(Pathogenic) | *KLHL40* | NM_152393.4:<br>c.1516A>C<br>p.(Thr506Pro)<br>(Pathogenic) | Missense/<br>AR | Nemaline myopathy 8<br>(IUGR, microcephaly,<br>multiple contractures,<br>ventilator dependent) | Unifying<br>monogenic<br>diagnosis achieved | 7 |
| 2 | ND | NcV | ND | NcV | ND | Neonatal alloimmune<br>liver disease - non-genetic (confirmed on<br>post-mortem<br>examination) | No unifying<br>monogenic<br>diagnosis.<br>Final diagnosis<br>achieved through<br>PM (non-genetic) | 6 |
| 3 | *VIPAS39* | NM_001400335.1:<br>c.1048-1G>A<br>(Pathogenic) | *VIPAS39* | NM_001400335.1:<br>c.1048-1G>A<br>(Pathogenic) | Splice site/<br>AR | *VIPAS39*-related ARCS2<br>(Arthrogryposis, renal<br>dysfunction, and<br>cholestasis) | Unifying<br>monogenic<br>diagnosis achieved | 5 |
| 4 | *FAM111A* | NM_001374866.1:<br>c.1451C>A<br>p.(Ala484Asp)<br>(VUS-3A) | *FAM111A* | NM_001374866.1:<br>c.1451C>A<br>p.(Ala484Asp)<br>(VUS-3A) | Missense/<br>*de novo*<br>AD | *FAM111A*-related<br>cranioosteostenosis<br>(DCDA twin, abnormally<br>shaped and thin cranium,<br>slender long bones,<br>fracture, platyspondyly<br>and hypoplastic ilia,<br>hydrops fetalis, ventilator<br>dependent) | Unifying<br>monogenic<br>diagnosis achieved | 7 |
| 5 | *NLRP1* | NM_033004.4:<br>c.3641C>G<br>p.(Pro1214Arg)<br>(Pathogenic) | *NLRP1* | NM_033004.4:<br>c.3641C>G<br>p.(Pro1214Arg)<br>(Pathogenic) | Missense/<br>*de novo*<br>AD | Autoinflammation with<br>arthritis and dyskeratosis<br>(severe CLD of uncertain<br>aetiology,<br>thrombocytopaenia<br>possibly related to NAIT,<br>conjugated hyperbilirubinemia with raised transaminases, fibrotic pulmonary vein stenosis) | Pathogenic NLRP1<br>may be<br>contributory but<br>unclear if other<br>monogenic factors<br>involved | 4 |
| **6** | *PTPN11* | NM_002834.5: c.1492C>T p.Arg498Trp (Pathogenic) | *PTPN11* | (NM_002834.5): c.1492C>T p.Arg498Trp (Pathogenic) | Missense/<br>*de novo*<br>AD | Noonan syndrome 1 (IUGR, complex congenital heart defect, including DORV, septal defects, mod PS, bilateral congenital diaphragmatic hernia, renal pelvis dilatation) | Unifying monogenic diagnosis achieved | 4 |
| **7** | ND | NcV | ND | NcV | ND | No unifying monogenic diagnosis (complex congenital heart defects, persistently raised methionine and homocysteine on NBS) | No unifying monogenic diagnosis for phenotype. Data reanalysis indicated and planned. | 6 |
| **8** | ND | NcV | ND | NcV | ND | No unifying monogenic diagnosis, DDx includes Fanconi anaemia and VACTERL (complex CHD – hypoplastic arch with coarctation and VSD, HLHS), duodenal atresia, sacral segmentation anomaly, anorectal malformation) | No unifying monogenic diagnosis, no pathogenic variants in genes associated with Fanconi anaemia | 5 |
| **9** | ND | NcV | ND | NcV | ND | No monogenic cause found for clinically unexplained neonatal liver failure and associated complication from disseminated intravascular coagulopathy) | No monogenic diagnosis for phenotype. Data reanalysis indicated and planned | 4 |
| **10** | *PCSK1* | NM_000439.5:<br>c.928G>A<br>p.(Gly310Arg)<br>(VUS-3A) | *PCSK1* | NM_000439.5:<br>c.928G>A<br>p.(Gly310Arg)<br>(VUS-3A) | Missense/<br>AR | *PCSK1*-related neonatal<br>severe generalized<br>malabsorptive diarrhoea<br>and failure to thrive<br>(profuse diarrhoea,<br>faltering growth, no<br>endocrinopathy) | Unifying<br>monogenic<br>diagnosis achieved | 4 |

NBG: Newborn Genomic Programme; VCGS: Victorian Clinical Genetic Services; AR: autosomal recessive; AD: autosomal dominant Solved: a clinical diagnosis and therapeutic management consistent with the HPO was made and implemented for the patient. ND; not determined; NcV, no clinically relevant variant; PM: post-mortem. Age of the probands at consent was between 1 and 60 days. \* TAT is working days only and excludes weekends and public holidays

## Discussion

Rapid genomic diagnosis offers potential benefits for critically sick patients that include guiding clinical management and improving prognosis.^16,30,49^ Although long-read sequencing technology is opening new opportunities for rapid diagnosis and treatment of rare genetic disorders, its adoption in clinical settings has been limited. This is despite evidence suggesting that integrating long-reads increases genetic testing capabilities beyond SNVs and small indels to include SVs, CNVs, and STRs^13–22^, as well as the possibility of moving testing to be closer to point-of care.^50^ The underutilization of this technology in the acute care of patients admitted in neonatal and paediatric intensive care units represents a gap, given the potential benefits it could offer for patient care and disease management.

The incorporation of nanopore sequencing into clinical practice holds significant potential. However, challenges exist regarding the stability and performance of LRS technology.^12,23,51–52^ To address these concerns, we have extensively benchmarked and validated our pipeline, emphasizing the importance of ensuring consistent and stable performance of the platform for successful routine implementation. Moreover, in line with clinical and technical recommendations,^45,53–54^ we have developed standardized protocols and quality control measures for long-read data to facilitate the integration of nanopore sequencing into clinical practice. These protocols ensure data consistency and reliability across different laboratory personnel, analytical teams, and platforms. Implementing such standardization measures is crucial for enabling the broader adoption of LRS technology in clinical settings.

Our established acute care clinical workflow integrates a highly scalable genome sequencing platform (*i.e.* PromethION 2 solo) with an AI-driven genomics analysis and interpretation software (Fabric GEM™; https://fabricgenomics.com/) to facilitate precise genome annotation, rapid variant interpretation, and prioritization. This pipeline has been established to provide clinicians with actionable information regarding SNVs, small indels, and SVs. In the initial benchmarking phase, we employed GA4GH benchmarking tools and GIAB samples sequenced to >30X coverage, demonstrating highly accurate variant calling metrics, particularly in high-confidence and coding regions of the human genome. Additionally, our workflow accurately detected large-scale CNVs in all eight samples from the Coriell Copy Number Variation Reference Panel 1 (CNVPANEL01), showcasing the capability of genotyping STRs and reliably profiling DNA methylation genome-wide.

Finally, in the validation phase, the clinical utility of long-read trio sequencing was demonstrated through the concordance of genomic findings with an established Acute Care Genomics service (VCGS; Melbourne, Australia), which utilized Illumina short-read sequencing technology.^43^ Notably, this concordance was achieved in all ten acute cases examined. These findings reinforce the potential of long-read sequencing for comprehensive genomic analysis and its applicability in clinical diagnostics.

The implementation of nanopore sequencing in the clinical care pathway is crucial.^16–18,47^ This study demonstrates the feasibility of such a pathway with the availability of funding, technology, skilled laboratory personnel, and researchers supporting rapid genomic testing for critically ill patients. The specialist multidisciplinary team (MDT) model is ideal for complex cases and provides clinicians with input for rare diagnoses that often lack established clinical management guidelines.^55,56^ In the evaluation of the cohort reported in this study, some cases remained undiagnosed, with at least one being non-genetic. In one case, the diagnosis of biallelic *PCSK1* congenital diarrhoea^57^ informed management and surveillance, with the family being reassured about the self-limiting nature of the diarrhoea and surveillance initiated through paediatric endocrine services. Three families used the results for reproductive risk assessment, while reanalysis of the negative trios will continue in the research setting. As expected in critically ill patients, many babies died. However, obtaining a diagnosis offers closure, enabling families to plan subsequent pregnancies.

In conclusion, we have successfully implemented a scalable clinical pipeline for rapid trio long-read whole-genome sequencing in an acute care setting, aiming to provide prompt and actionable genomic information to clinicians. Through this effort, we have demonstrated the feasibility of achieving rapid precision medicine for critically sick children on a national scale using long-read technology.

## Data Availability

The genomic data (i.e. bam files) for the Genome in a Bottle (GIAB) samples is publicly available from NCBI Sequence Read Archive (SRA). The genomic and phenotypic data from families analysed in this study cannot be shared publicly due to privacy and ethical restrictions.

## Acknowledgements

The authors would like to express our thanks to the families who participated in the sequencing programme during times of great stress. The authors would like to thank: Anita Lee and the Genetic Counsellors and Clinical Geneticists at Genetics Health Services New Zealand; the Neonatologists, Paediatricians, Clinical specialists, nurses and support staff at the Neonatal Intensive Care and Paediatric Intensive Care Units at Auckland City Hospital; the staff at Diagnostic Genetics, LabPLUS for their discussions, and the clinical care of the patients involved in this study.

We gratefully acknowledge the generous donations from the Dines Family Trust, The Toutoku Trust, and The Kelliher Trust.

The authors wish to acknowledge the use of New Zealand eScience Infrastructure (NeSI) high performance computing facilities, consulting support and/or training services as part of this research. New Zealand’s national facilities are provided by NeSI and funded jointly by NeSI’s collaborator institutions and through the Ministry of Business, Innovation & Employment’s Research Infrastructure programme (https://www.nesi.org.nz).

## Author Contributions

Conceptualization: J.M.O.; Study design: J.M.O., D.M.N., P.T., C.G., H.H.P., P.Q.S., S.F., G.T., E.T., K.G., J.V.D., P.Y., M.G; Bioinformatics analysis: D.M.N., P.T.; Writing-original draft: D.M.N., P.T.; Visualization: D.M.N., P.T.; Formal analysis: D.M.N., P.T.; Methodology: C.G., H.H.P., J.R., S.F; Ethics: C.G., J.V.D; Investigation: J.M.O., D.M.N., P.T., C.G., H.H.P., P.Q.S., S.F., J.V.D., P.Y., M.G; Funding acquisition: J.M.O; Resources: J.M.O., J.V.D; Patient recruitment: P.Y., K.G. Writing-review & editing: J.M.O., D.M.N., P.T., C.G., H.H.P., P.Q.S., S.F., G.T., E.T., S.L., Z.S., K.G., J.V.D., P.Y., M.G; Principal investigator: J.M.O. All authors approved the submission of this manuscript.

These authors contributed equally D.M.N., PT., CG., H.H.P., P.Y.

## Ethics Statement

Ethics approval was obtained from the Northern B Health and Disability Ethics Committee for the study entitled: *Newborn Genomics – Te Ira oo Te Arai* (Ethics reference: 2023 FULL 15542). Locality approval was obtained from the Research Review Committee Te Toka Tumai Auckland for the project entitled: *Newborn Genomics – Te Ira oo Te Arai* (Reference A+9855 [FULL 15542]).

## Patient Consent Statement

Parents of the participating newborns provided written informed consent.

## Data Availability

The genomic data (*i.e.* bam files) for the Genome in a Bottle (GIAB) samples is publicly available from NCBI Sequence Read Archive (SRA) under BioProject accession number: PRJNA1117929. The genomic and phenotypic data from families analysed in this study cannot be shared publicly due to privacy and ethical restrictions.

## Code Availability

Pipelines and software used for the analyses reported in this manuscript are publicly available. EPI2ME workflows for aligning FASTQ files sequences to the reference genome is available on https://github.com/epi2me-labs/wf-alignment and the human variation pipeline for variant calling (*i.e.* SNVs/indels, SVs, CNVs, STRs), and modified bases analysis can be accessed on https://github.com/epi2me-labs/wf-human-variation. Hap.py is available on https://github.com/Illumina/hap.py and Truvari on https://github.com/ACEnglish/truvari. Dorado is available on https://github.com/nanoporetech/dorado. Modkit is available on https://github.com/nanoporetech/modkit. Samtools is available on https://github.com/samtools/samtools. Modbamtools is available on https://github.com/rrazaghi/modbamtools. A docker image containing seaborn, matplotlib, and python libraries used to generate plots is available as docker://nyagam/seaborn:latest.

## Supplementary Information

Supplementary files, figures and tables are available in the Supplementary documents section.

