## Supplementary Figure for "Benchmarking and quality control for nanopore sequencing and feasibility of rapid genomics in New Zealand: validation phase at a single quaternary hospital"

**Benchmarking and quality control for clinical nanopore sequencing facility for acute care in New Zealand**

^7^LabPLUS, Auckland District Health Board

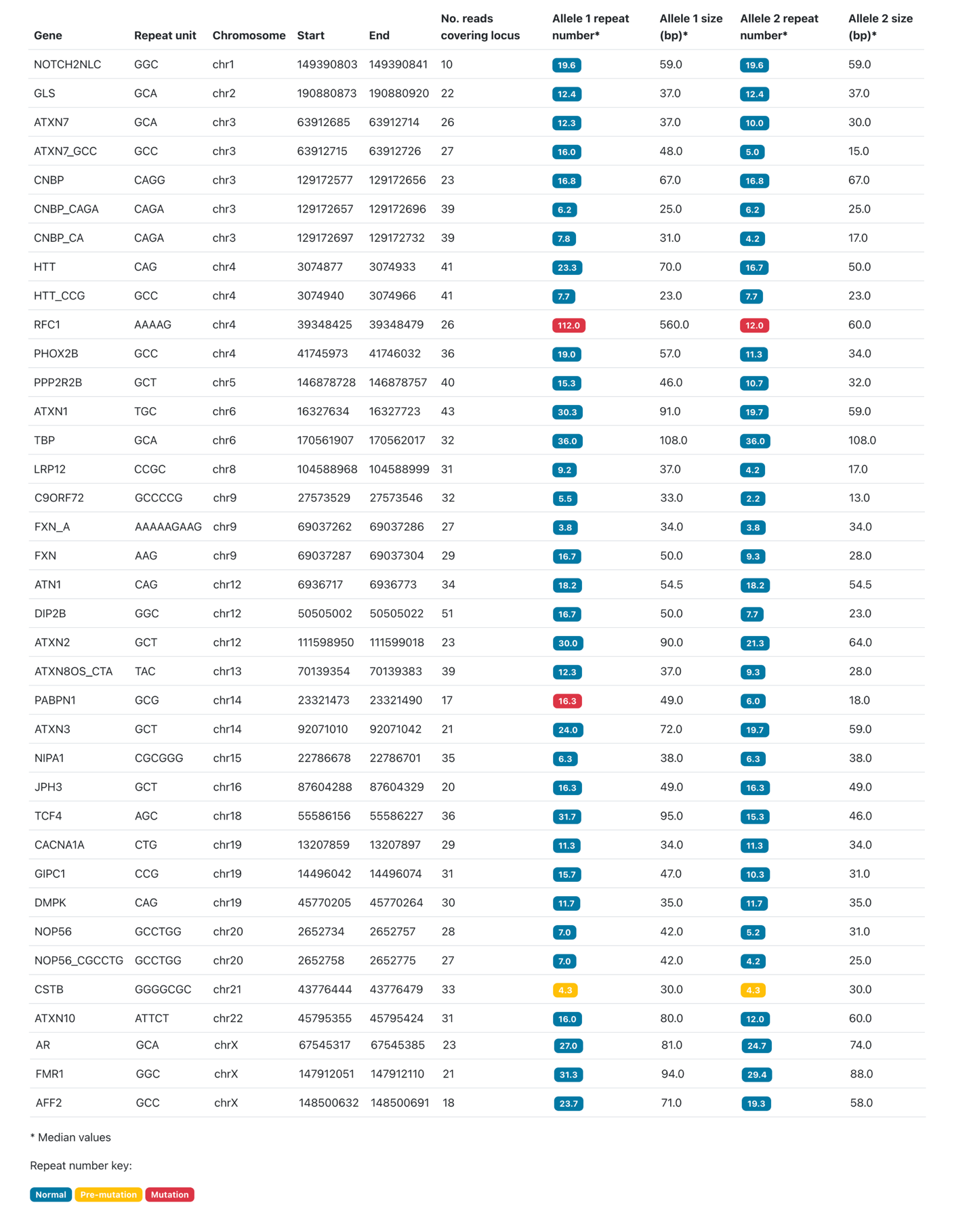

**Supplementary Figure 1. A summary of the repeat expansions genotyped and quantified in the HG002 genome.** ONT reads are capable of genotyping 37 clinically relevant genomic regions associated with repeat expansions using Straglr (https://github.com/philres/straglr) as implemented in EPI2ME Labs’ wf-human-variation pipeline v1.7.0.

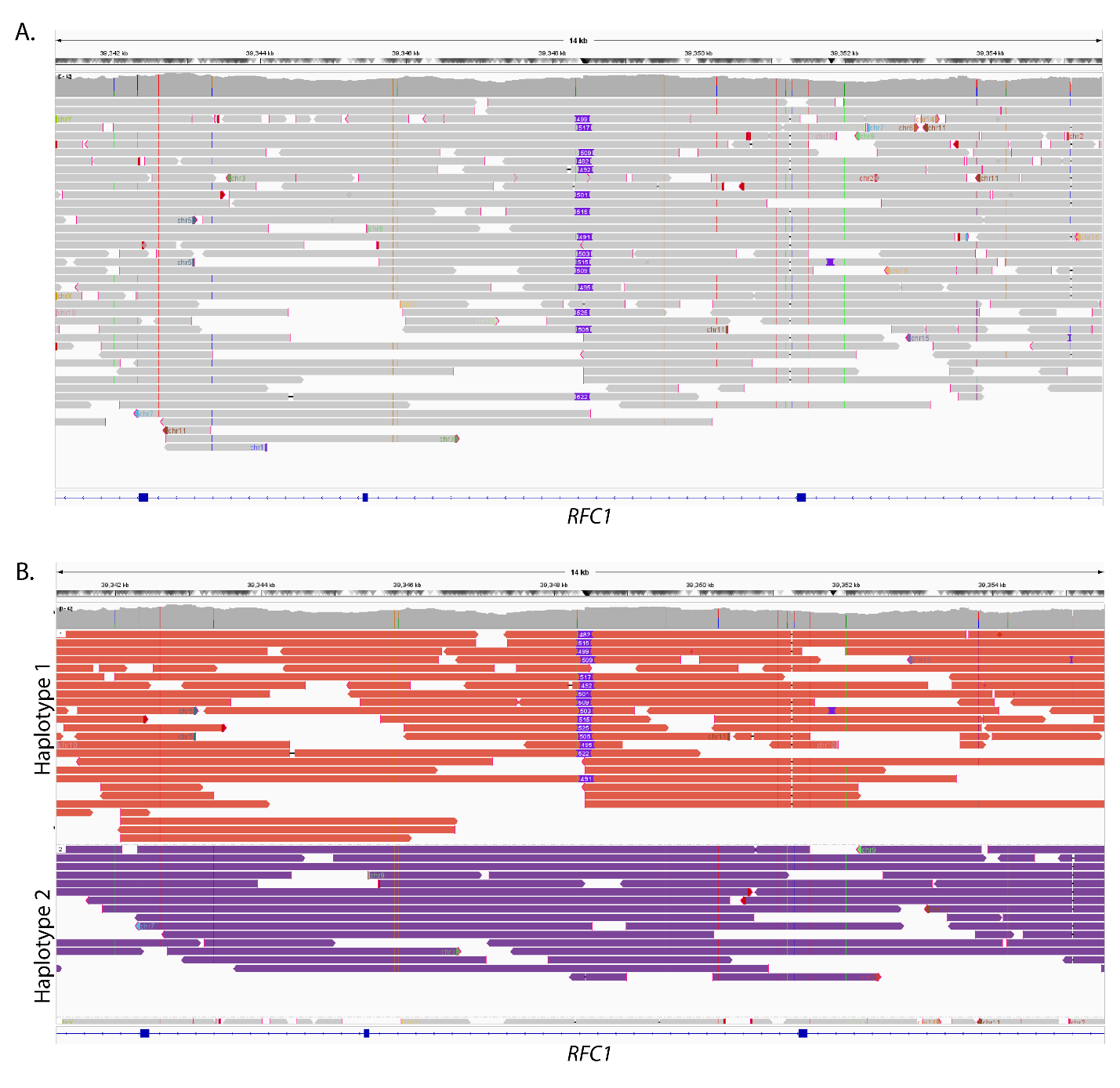

**Supplementary Figure 2.** Integrative Genomic Viewer visualization of: **A)** the genotyped *RFC1* repeat expansion represented as a ~500bp heterozygous insertion (purple box), and **B)** haplotype-specific information of the insertion, which indicates that the repeat elements are mapped on haplotype 1 (orange reads [top]) and not haplotype 2 (purple reads [bottom]).

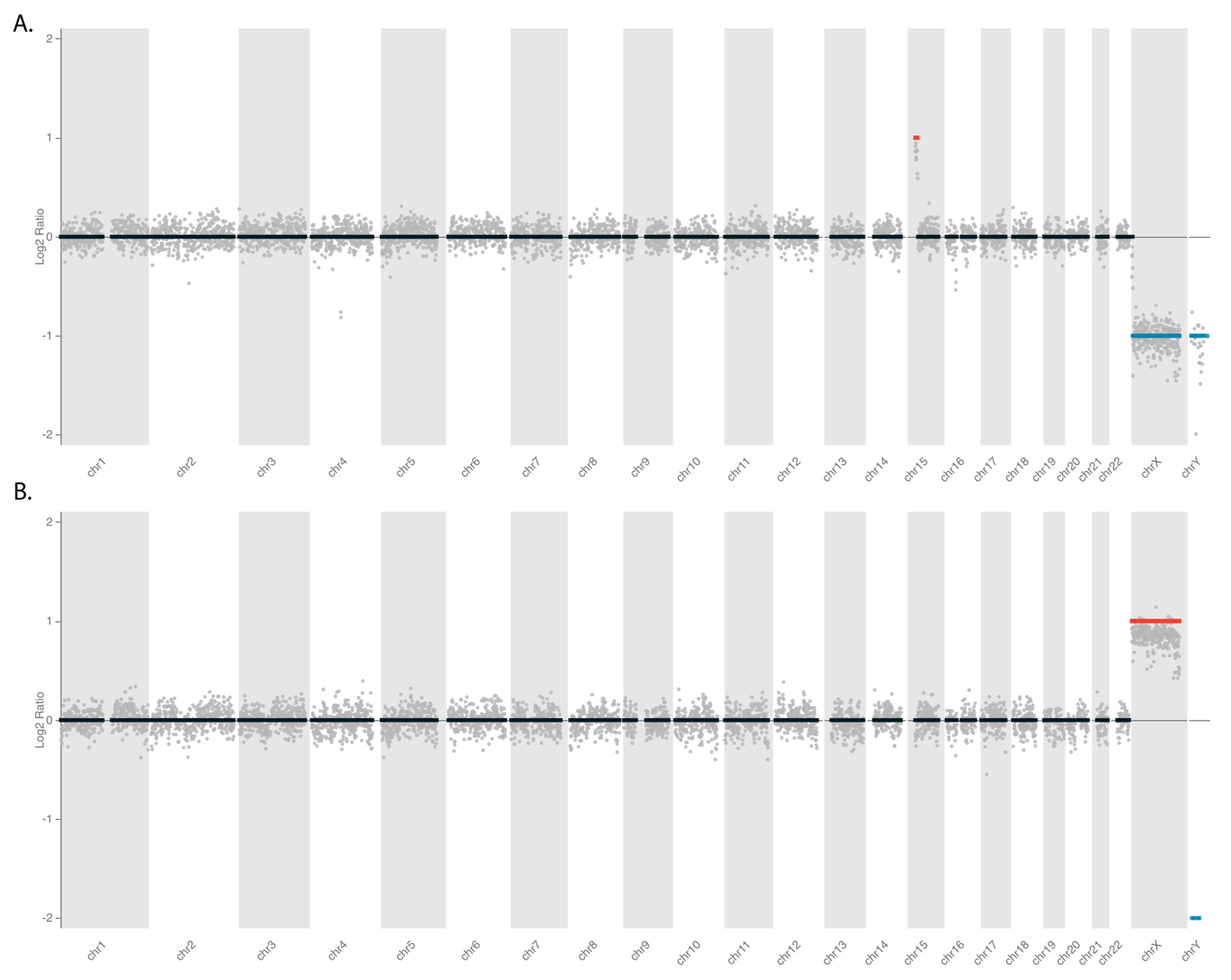

**Supplementary Figure 3.** Smoothed read counts for copy number profiles of **A)** GM20556 (the isodicentric chromosome 15 CNV) and (B) GM01416 (XXXX syndrome CNV) genomes at ~2X coverage, corresponding to ~2.6M reads. Bins (500Kbp) are ordered along the x-axis by their genomic positions, and the y-axis shows median-normalized log 2 -transformed data. Red color: increased copy number detected; Blue: decreased copy number detected; Black: read counts of < -2 converted to 2 (i.e. 2 copies of each chromosomal segment).

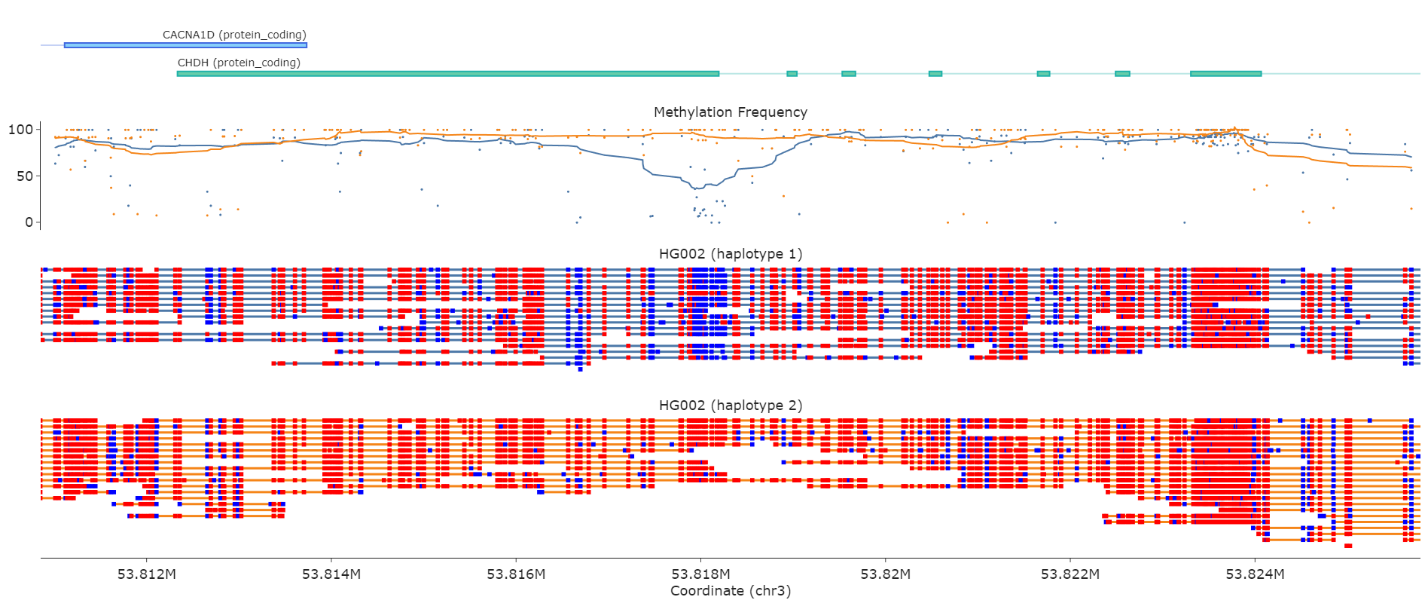

**Supplementary Figure 4**. A novel haplotype-specific differential methylation in *CHDH* as visualized using modbamtools (https://github.com/rrazaghi/modbamtools). Methylated CpG sites are denoted in red, while unmethylated sites are represented in blue. A methylation frequency plot and gene locus are visualized above the haplotype reads.
