## Supplementary File 1 for "Benchmarking and quality control for nanopore sequencing and feasibility of rapid genomics in New Zealand: validation phase at a single quaternary hospital"

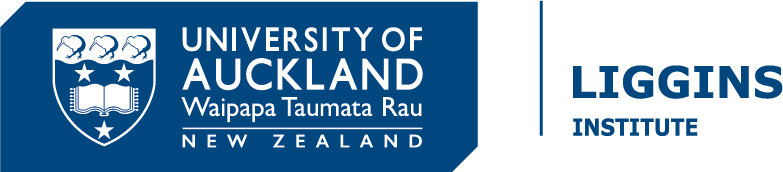

Participant Information Sheet

Newborn Genomics Programme

Te Ira ō te Arai | Beyond the veil

### Trio Whole Genome Sequencing for investigating genetic causes of rare disease

| Project Leader | Patrick Yap |
| --- | --- |
| Address | Genetic Health Service NZ, Building 30, Auckland City Hospital |
| Contact phone number  Ethics committee ref: | 09-307-4949 ext 25959  2023 FULL 15542 |

### Introduction

You are invited to take part in a research study called the “Newborn Genomics Programme - Te Ira ō te Arai” which is being carried out in partnership between Te Toka Tumai, Auckland City Hospital and The Liggins Institute at the University of Auckland.

You are invited to take part in this study because your child may have a rare genetic condition. Participation in this study is voluntary, your choice. If you do not want to take part, you do not have to give a reason, and it will not affect the care your child receives. If you do want to take part now, but change your mind later, you can pull out of the study at any time.

This information sheet will help you decide if you would like to take part. It explains why we are doing the study, what your participation involves, and the benefits and risks. We will go through this information with you and answer any questions you may have. Before any decision, you may want to talk with other people, such as whānau, friends, or healthcare providers. Feel free to do this.

If you agree to take part in this study, you will be asked to sign the Consent Form. You will receive a copy of the Information Sheet and the Consent Form to keep.

This document is 7 pages long, excluding the Consent Form. Please make sure you have read and understood all the pages.

### What is the study about?

The aim of this study is to identify the cause of your child’s condition. We are using a new type of genetic test that allows us to map out the sequence (spelling) of your and your child’s DNA (“whole genome sequencing”). We want to understand how useful this new type of test is and discover new genes that cause rare genetic syndromes.

### What does “whole genome sequencing” mean?

Genes are pieces of DNA that contain the instructions for how your body works. We all have thousands of genes inside every cell in our body. Traditional genetic testing can only look at one gene at a time. “Whole genome sequencing” lets us look at all of your genes at the same time. This means it is a fast way to search for disease-causing changes inside and outside of genes, which we call “variants”.

Every person has millions of variants in and around their genes. This makes it hard to work out which variants are harmless, and which cause disease. This study will compare your child's DNA to your DNA to help identify which variants might be associated with their health problems. This approach is known as “Trio whole genome sequencing”.

### What would participation involve?

1. We will ask you for your informed consent. Before you sign the Consent Form, you will receive genetic counselling during which the study benefits and risks will be explained to you and you will be able to ask any questions. You will receive a copy of the Participant Information Sheet and Consent Form to keep.

2. We will access your and your baby’s medical information collected by your specialists. Only the researchers involved in this study will have access to your medical notes. To help confirm genetic findings, further tests might be suggested to your treating doctor, such as more blood tests or imaging. Your doctor would discuss these with you.

3. DNA collection: For this study we will need a sample of genetic material (DNA) from your affected child, and both parents. We will ask you to provide two blood samples. We will take one blood and one saliva sample from your child. DNA and RNA will be extracted from these samples.

### What will happen to my samples?

Sample preparation and storage

Your samples will be sent to the Diagnostic Genetic Laboratory at LabPlus, a clinically accredited laboratory. LabPlus will retain a blood sample and your child’s saliva sample. These samples will be registered under your name and personal information. LabPlus will extract DNA and RNA from your and your child’s blood and saliva samples for concurrent clinical testing. The remaining DNA will be stored in the clinical laboratory, following national laboratory guidelines.

The remaining blood samples of you and your child will be sent to The Liggins Institute. Before sending the blood samples for sequencing, the research team will de-identify your samples by replacing your and your baby’s name with a code. This code is randomly generated by the computer. This will protect your privacy. Only the study investigators who lead the project can link the code to your identity.

We will extract DNA and RNA from your and your child's blood and saliva samples in the research laboratory at The Liggins Institute. The remaining DNA will be stored in the research laboratory, following national laboratory guidelines.

Sending samples for sequencing

The de-identified DNA from you and your child will be sequenced at the Liggins Institute. Your sequenced DNA data will be stored on a dedicated secure research storage system.

Genome sequencing enables researchers to determine the DNA sequence of our whole genome. Your DNA sequence can then be analysed to determine if you have any specific changes that put you at risk of a genetic disease.

In addition to sequencing at The Liggins Institute your de-identified DNA extracted from your blood samples at The Liggins Institute may be sent to Victorian Clinical Genetics Services, Melbourne, Australia for sequencing and analysis. Victorian Clinical Genetics Services is a well-established laboratory in Melbourne that has been providing clinical-grade genomic testing. Sending DNA to Victorian Clinical Genetics Services lets us check the results that we obtain from our method for reassurance.

Only study staff at the Victorian Clinical Genetics Services will have access to your DNA sample and sequence data. Your DNA sample will not be retained at the Victorian Clinical Genetics Services.

You do not have to agree to send your samples to the Victorian Clinical Genetics Services. You may opt out of this. If you opt out your samples will still be analysed in Auckland. This will not affect you or your child’s future health care. If you do opt in to having your samples sent to Victorian Clinical Genetics Services for analysis the process will be explained to you, and you will be asked to sign an additional consent form.

*Analysis of your sequenced DNA*

After sequencing your de-identified DNA sequencing data will be analysed locally and using cloud computing services which are based overseas to identify which ones might be responsible for their condition. Your and your child’s DNA data will not be retained by this software.

Sample Biobanking/Disposal

After sequencing, any remaining blood samples will be transferred to The Auckland Regional Biobank - Te Ira Kawai. Submitting your samples to The Auckland Regional Biobank – Te Ira Kawai is mandatory.

The purpose of the databank / registry is to store your de-identified blood samples to ensure that the DNA sequence can be regenerated as needed. They will not be used for any other studies and will be destroyed after 26 years unless you opt to have your samples returned to you. Samples will be returned to you from the biobank after they have been stored for 26 years.

Alternatively, if you wish, the unused samples can be disposed of with a karakia (blessing). You can choose how the samples are disposed of in the consent form.

### What will happen to the genetic information found during the study?

Genetic information obtained about you or your child during this study is securely stored. No information will be released to any outside people or groups. This includes insurance companies, employers, or the police, with the exceptions below.

We may share genetic data, in a way that does not identify you, with other approved research teams. The information will remain de-identified. We may do this to increase the chances of making important discoveries about a genetic condition. For example, comparing your DNA to DNA from other unrelated people with the same condition would help us understand the condition. We would let your clinician know any new information arising from these studies.

De-identified genetic data is sometimes added to controlled access databases (e.g. the United States National Institutes of Health (NIH) database of Genotypes and Phenotypes (dbGaP)). Safeguards are in place to protect your information while it is stored in these databases and used for research. Researchers who wish to access these databases are evaluated by a central data access committee, which awards access if the proposed usage is ethical and responsible. The data will be sent with only a code number attached. Your name, and other identifiable information, will not be provided. This would only happen if comparing your genetic data with that from thousands of people would help to improve our understanding of the changes we found in your DNA. You may not directly benefit from this work, but your information would help advance knowledge for everyone. Your personal details will not be provided to researchers.

You may hold beliefs about sacred and shared values of any tissue samples removed and data originating from the tissue. The cultural issues associated with sending your tissue samples and data overseas and/or storing your tissue and data should be discussed with your family/whanau as appropriate. If you need cultural support this can be provided. Please let us know and we will arrange this for you, or you can ring the number at the bottom of the participant information and consent form. Cultural support is different to knowing more about the study treatments. In these cases, we can arrange a primary investigator to come and talk to you and your whānau.

In co-developing this programme, we have consulted with the Liggins Māori Advisory Committee. In taking samples we recognize the takoha – gift of responsibility that accompanies the samples, which for many are tapu and imbued with wairua, and undertake to return the results and future outcomes to the whanau so that they can add to their whakapapa (past, present, and future).

Options for karakia will be discussed with you during the informed consent process if you wish.

Māori Data Sovereignty

*Māori data sovereignty* is about protecting information or knowledge that is about (or comes from) Māori people. We recognise the taonga of the data collected for this study. To help protect this taonga:

- We have consulted with the Liggins Māori Advisory Committee about the collection, ownership, and use of study data.
- Māori data sovereignty principles will be applied to all Māori data that is produced from biological samples

### Will I be given the results of the study?

Any research results that could be significant to your child or your family will be confirmed either by an accredited diagnostic laboratory like Victorian Clinical Genetics Services independently or by using a different method that has been proven to be reliable. This is standard practice for genetic testing. These tests would be provided free of charge to you and your child.

If we find the cause of your child’s condition, your specialist will discuss these results with you. Sometimes we find results that are important for you as well. It is important to read the information under ‘risks’ so you can decide if you wish to be told the test results.

### Are there any benefits of participating in the study?

This research study may identify a change in your child's DNA that explains their condition. There is published evidence indicating that some sick newborns may benefit from rapid diagnosis (within a week of testing). A definitive diagnosis may help target and/or fine-tune medical management. The data analysis will initially focus on finding a diagnosis based on your child’s clinical presentation. The results will be discussed with you through the referring specialist and Clinical Geneticist/Genetic Counsellor within a week.

In situations where the testing does not provide a definitive diagnosis, data analysis will continue to occur at a regular interval (usually every 12months) with newly available evidence from research and publications. Repeat analysis will only examine your data for changes in your child’s DNA that may explain their condition. Your data will not be used for broader, unspecified research. If you would like to receive this information, we will forward the results to your doctor. Genetic testing may also be possible for your wider family. This information may also help understand your health, and the risks of the same change occurring in another pregnancy. This study will improve our understanding of the genetic basis of rare conditions and benefit all people with these conditions.

It is possible that there is no direct benefit from this research for you or your family. This form of genetic testing uses “state of the art” technology. However, we still cannot find the cause of disease in all affected individuals.

### Are there any side-effects and risks associated with this study?

We will ask you to give a blood sample. We will take a blood and saliva sample from your child. We expect no other problems in taking part in this study.

Learning the genetic test results for your child might create uncertainty or may be upsetting for you. For example, if the disease has no known treatment or cure. Genetic counsellors and doctors will support you if this happens.

Another risk relates to the chance of making an unexpected or “incidental” inheritable genetic finding. For example, we could discover that you or your child have an increased risk of another health condition. To reduce the chance of making these incidental findings, we only look at the genetic changes that are relevant to your child’s condition. We will not screen for other inherited conditions. It is still possible that we will discover something unrelated, that is relevant to your health, or your family’s health. It is also possible that we will identify instances of misattributed parentage. Misattributed parentage occurs when genetic testing reveals that either the father, the mother or both parents are not the biological parent. Worldwide, other researchers have found that the chances of making these incidental findings are low and happens in less than 1% of cases.

If testing reveals an incidental finding, it is up to you whether you would like to receive further information. You can opt out of receiving any information about incidental findings. If you wish to find out about incidental findings that are important for your health, the information will be passed to your referring specialist to arrange appropriate follow-up care and/or genetic counselling.

You might know that your family has a high risk of carrying a genetic change that increases your risk of developing adult-onset conditions and cancer risk. If you would like us to explore these genetic changes, please talk to your specialist. It is possible to examine these risk factors ONLY if there is significant family history. This can be determined by the specialists (Geneticists and Genetic Counsellors) working at the Genetic Health Service New Zealand. We can NOT perform analysis of these types of risk factors in minors (under the age of 18 years). We do this to protect the minor’s autonomy to make their own decisions to understand this risk when they are adults.

Some people worry that their genetic information gets into the wrong hands and is used in the future to discriminate against them. We think that this is very unlikely to occur because of the care we will take to keep this information confidential.

We might find something of major significance for your health and report this back to you. If this happens, private insurers would consider this information as “prior knowledge” of a medical condition. Statutory or contractual duties may require you to disclose these results to third parties, for example, insurance companies, employers, and financial and educational institutions.

If you were injured in this study, which is unlikely, you would be eligible to apply for compensation from ACC. This is just as you would be if you were injured in an accident at work or at home. This does not mean that your claim will automatically be accepted. You will have to lodge a claim with ACC, which may take some time to assess. If your claim is accepted, you will receive funding to assist in your recovery. If you have private health or life insurance, you may wish to check with your insurer that taking part in this study won’t affect your cover.

### What are the rights of participants in the study?

Freedom of Consent:

This is a voluntary study. Whether or not you take part is your choice. You are invited to discuss the study with your family/whanau.

If you do agree to take part, you are free to withdraw from the study at any time. This will not affect you or your child’s future health care. You can withdraw your DNA samples from the study at any time prior to testing. If the DNA sample is sent to an overseas laboratory this will be destroyed and not be returned. If you withdraw from the study, all of your New Zealand based samples will be destroyed by standard lab disposal methods. All tests and procedures will be free of charge.

Ethics require we keep samples on children for 26 years in case we need to check anything. After 26 years all of the New Zealand samples you provided to this study will be destroyed unless you opt to have your samples returned to you. Samples will be returned to you from the biobank after they have been stored for 26 years.

You may believe that the tissue samples needed for this study are sacred or have a shared value. Cultural issues associated with sending your samples overseas and/or storing your tissue should be discussed with your family/whānau as appropriate. There are a range of views held by Māori around these issues; some iwi disagree with the storage of samples and advise their people to check with their iwi prior to participation in research where this occurs. However, it is acknowledged that individuals have the right to choose.

Reconsent of participants

All participant blood samples will be retained at The Auckland Biobank – Te Ira Kawai for 26 years as per ethics requirements.

You will be providing consent for your child to participate in this study and therefore have their samples also stored for 26 years. Once your child turns 16, they can decide for themselves whether they want to consent (agree) to having their blood samples retained for a further 10 years.

Your child will be followed up at age 16 to confirm if they wish to re-consent (agree) to their involvement in the research study.

### Confidentiality:

All information collected as part of this study will be kept confidential and stored securely. Genetic material/samples that are sent overseas will be identifiable only by a code. Only the principal investigator will be able to match the code and participant. Once all personal identification is removed, results of the study may be presented in public talks or written articles. No information will be shown that identifies your child without your written permission to do so.

### Statement of Approval

This study has received ethical approval from Northern B Health and Disability Ethics Committee.

### Where can you go for more information about the study, or to raise concerns or complaints?

If you have any questions, concerns, or complaints about the study, you can contact:

Name: Patrick Yap, Clinical Geneticist

Telephone number: 09-307-4949 ext 25959

If you want to talk to someone who isn’t involved with the study, you can contact an independent health and disability advocate on:

Fax : 0800 2 SUPPORT (0800 2787 7678)

If you require Māori cultural support, talk to your whānau in the first instance. Alternatively, you may contact the administrator for He Kamaka Waiora (Māori Health Team) by telephoning 09 486 8324 ext 43553

You can also contact the health and disability ethics committee (HDEC) that approved this study on:
