## Supplementary File 2 for "Benchmarking and quality control for nanopore sequencing and feasibility of rapid genomics in New Zealand: validation phase at a single quaternary hospital"

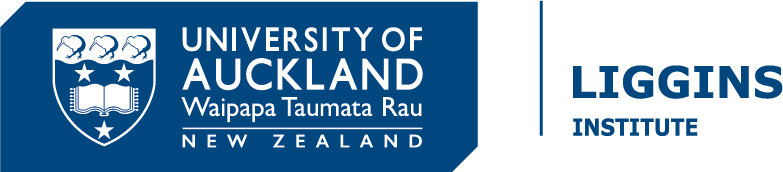

CONSENT FORM

### Newborn Genomics Programme: Trio whole genome sequencing

Project Leader	:	Patrick Yap, Clinical Geneticist

Address:	Genetic Health Service NZ, Building 30, Auckland City Hospital

Contact phone number:	09-307-4949 ext 25959

Ethics committee ref.:	2023 FULL 15542

#### **Full names of researchers**:

Patrick Yap, Clinical Geneticist, Genetic Health Service NZ, Auckland City Hospital

Justin O’Sullivan, Professor of Genetics and Systems biology, Liggins Institute, University of Auckland

Contact phone number: 09-307-4949 ext 25959 (Patrick Yap)

| Participation in the study | Please initial |
| --- | --- |
| I have read and understood the information sheet about this study, and I understand what is involved. |  |
| I understand that I will be given a copy of this Consent Form and the Information Sheet to keep. |  |
| I have been given the opportunity to discuss this study and to ask questions about it. I am satisfied with the answers I have been given. |  |
| I have had enough time to consider whether my child and I take part, and to discuss my decision with a person of my choice including whānau/family members if I wish. |  |
| I know who to contact if I have questions about the study. |  |
| I understand that taking part is voluntary and I am free to withdraw my child or myself from this study at any time and for any reason. |  |
| I understand that my child’s and my participation in this study is confidential and no information that could identify us will be used in any reports on this study. |  |

| Use of my child’s and my samples and medical information | Initial or choose options |
| --- | --- |
| I consent for the researchers to access my child’s and my medical notes. |  |
| I consent for my child and myself to provide blood and saliva samples for this study if requested. |  |
| I am aware that the study will store and analyse my child’s and my genetic material, and I consent for such analysis being performed. |  |
| I am aware that my and my child’s de-identified DNA sequencing data will be analysed locally AND using cloud computing services which are based overseas, and I consent for such analysis being performed. |  |
| I am aware that my child’s and my DNA may be sent overseas for sequencing. |  |
| I understand that the data generated by sequencing my child’s and my DNA will be shared with other researchers and stored on a database only after all personal identifiers such as name, address and date of birth have been removed. |  |
| I understand that ongoing reanalysis of the genetic information may be required in order to find a diagnosis. I consent to the reanalysis of my child’s and my genetic information and medical notes for up to ten years. |  |
| I consent for the samples to be stored for up to 26 years within The Auckland Regional Biobank. I understand that I can request to have my child’s and my samples returned or destroyed at any time. |  |

| Disclosure of results | Initial or choose options |
| --- | --- |
| I understand that if the study finds the genetic cause for my child’s current medical condition(s), I will receive results through a genetic counsellor or clinical geneticist. |  |
| I understand that the genetic testing may produce unexpected and unsought clinically actionable incidental findings that are NOT related to my baby’s disorder being studied (please see Participant Information Sheet for details). |  |
| I wish to be informed of clinically actionable incidental findings. | YES / NO |

| Future unspecified use of genetic information | Please initial |
| --- | --- |
| I agree that de-identified genetic information about my child and me may be kept in databases (in New Zealand or abroad) and used in future studies into genetic diseases. Please note that identifying information about you, such as your name, address, date of birth, telephone number, will **NOT** be put into any databases. |  |

| Remaining samples instructions | Please tick **ONE** option |
| --- | --- |
| I consent to any remaining samples(s) being disposed of using standard disposal methods at the end of this study. |  |
| I elect to have all my child’s and my remaining samples disposed of with an appropriate karakia at the end of this study. |  |
| I elect to have my child’s and my remaining samples returned to me at the end of this study. |  |

I, __________________________________ (print full name), hereby consent for my child ___________________________________ (print full name) and myself to take part in this study.

………………………………………………	………………	…………………………………………….

Name of Participant (you)	Date	Signature

………………………………………………	………………	…………………………………………….

Name of whānau/family member	Date	Signature

(optional)

I have given a verbal explanation of the research project to the participant and have answered the participant’s questions about it. I believe that the participant understands the study and has given informed consent to participate.

………………………………………………	………………	…………………………………………….

Project explained by (name)	Date	Signature
